# Chromosome 10q24.32 Variants Associate with Brain Arterial Diameters in Diverse Populations: A Genome-Wide Association Study

**DOI:** 10.1101/2023.01.31.23285251

**Authors:** Minghua Liu, Farid Khasiyev, Sanjeev Sariya, Antonio Spagnolo-Allende, Danurys L Sanchez, Howard Andrews, Qiong Yang, Alexa Beiser, Ye Qiao, Emy A Thomas, Jose Rafael Romero, Tatjana Rundek, Adam M Brickman, Jennifer J Manly, Mitchell SV Elkind, Sudha Seshadri, Christopher Chen, Ralph L Sacco, Saima Hilal, Bruce A Wasserman, Giuseppe Tosto, Myriam Fornage, Jose Gutierrez

## Abstract

**Background:** Brain arterial diameters are novel imaging biomarkers of cerebrovascular disease, cognitive decline and dementia. Traditional vascular risk factors have been associated with brain arterial diameters but whether there may be genetic determinants of brain arterial diameters is unknown.

**Results:** We studied 4150 participants from six geographically diverse population-based cohorts (40% European, 14% African, 22% Hispanic, 24% Asian ancestries). We measured brain arterial diameters for 13 segments and averaged them to obtain a global measure of brain arterial diameters as well as the posterior and anterior circulations. A genome-wide association study (GWAS) revealed 14 variants at one locus associated with global brain arterial diameter at genome-wide significance (P<5×10^−8^) (top SNP, rs7921574; β =0.06, P=1.54×10^−8^). This locus mapped to an intron of *CNNM2*. A trans-ancestry GWAS meta-analysis identified two more loci at *NT5C2* (rs10748839; P=2.54×10^−8^) and at *AS3MT* (rs10786721; P=4.97×10^−8^), associated with global brain arterial diameter. In addition, two SNPs co-localized with expression of *CNNM2* (rs7897654, β=0.12, P=6.17×10^−7^) and *AL356608*.*1* (rs10786719, β =-0.17, P=6.60×10^−6^) in brain tissue. For the posterior brain arterial diameter, two variants at one locus mapped to an intron of *TCF25* were identified (top SNP, rs35994878; β =0.11, P=2.94×10^−8^). For the anterior brain arterial diameter, one locus at *ADAP1* was identified in trans-ancestry genome-wide association analysis (rs34217249; P=3.11×10^−8^).

**Conclusion:** Our study reveals three novel risk loci (CNNM2, NT5C2 and AS3MT) associated with brain arterial diameters. Our finding may elucidate the mechanisms by which brain arterial diameters influence the risk of stroke and dementia.

## Background

Dolichoectasia has been defined by elongated and tortuous arteries ^1^ and it is usually associated with smoking, male sex and aging.^2^ The diagnosis of dolichoectasia has been historically ascertained by visual inspection of neuroimaging or more recently using a fixed arterial diameter cutoffs for the basilar artery.^3^ Although these methods are easy to use, they simplify the biological meaning of the continuum of intracranial arterial diameters in brain health and neglect arterial-size expectations based on age, sex and head size.^4^ Therefore, we proposed and validated the principle that arterial diameters measured continuously and adjusted for head size relate to health outcomes in a non-linear fashion and that people with very small or very large arterial diameters are at a higher risk of vascular events.^5^ Furthermore, dilated brain arterial diameters are associated with a higher risk of dementia ^6^ and steeper cognitive decline.^7^ Smaller arterial diameters causing stenosis, usually related to atherosclerosis, are intuitively related to adverse health outcomes ^8,9^ but less is known about the underlying nature of dilated brain arteries.

Larger arterial diameters have been described in people with connective tissue disorders such as Marfan syndrome,^10^ Ehlers-Danlos,^11^ and Arterial Tortuosity Syndrome,^12^ among others. These monogenic diseases are usually rare and detected in younger patients. Furthermore, there is clear association between larger arterial diameters and vascular risk factors, especially hypertension.^13,14^ Although hypertension is highly prevalent in elderly populations, the heterogeneity of brain arterial phenotypes in people with vascular risk factors suggests that a specific genetic profile might partially be responsible for higher risk of brain arterial dilatation. Consequently, we hypothesize that in the general population less pathogenic but more frequent genetic variants may relate to brain arterial diameters. Identifying such a genetic profile may shed light into possible mechanistic links between large brain arterial diameters and the observed brain outcomes. To test our hypothesis, we leveraged diverse population cohorts within and outside the United States to investigate associations between brain arterial diameters and Alzheimer’s disease, stroke, and white matter hyperintensities volume.

## Results

### Multi-ancestry GWAS Identifies a Novel Locus Associated with Brain Arterial Diameter

We conducted a multi-ancestry GWAS for brain arterial diameter levels in 4150 participants, including 1650 from European, 583 from African, 920 from Hispanic, and 997 from Asian ancestries. Mean age of the participants across studies ranged from 70 to 76 years, with proportions of women ranging from 52 to 64%. Detailed demographic information is presented in Table 1.

**Table 1.** Demographic information of studies

| Cohort | ARIC | NOMAS | WHICAP | EDIS | MCS | FHS |
| --- | --- | --- | --- | --- | --- | --- |
| N | 1565 | 1092 | 290 | 647 | 350 | 206 |
| Age, years, mean (sd) | 75.8 (5.3) | 70.1 (8.4) | 77.2 (6.5) | 70.2 (6.6) | 71.0 (8.2) | 72.7 (11.7) |
| Female (%) | 58.8 | 60.5 | 64.1 | 51.7 | 55.7 | 54.3 |
| Self-reported racial/ethnic identity or geographic ancestry (%) |  |  |  |  |  |  |
| European/white | 74.3 | 13.8 | 44.8 | - | - | 100.0 |
| African/black/African American | 25.7 | 16.6 | 55.2 | - | - | - |
| Hispanic | - | 69.6 | - | - | - | - |
| Asian | - | - | - | 100.0 | 100.0 | - |
| Hypertension (%) | 74.9 | 68.1 | 63.1 | 80.8 | 67.8 | 68.4 |
| Diabetes (%) | 33.9 | 19.0 | 20.2 | 37.3 | 31.8 | 23.9 |
| Dyslipidemia (%) | 56.0 | 45.0 | 33.9 | 76.0 | 71.4 | 51.6 |
| Smoking (%) | 6.0 | 52.7 | 5.2 | 27.9 | 7.1 | 9.3 |

ARIC, Atherosclerosis Risk in Communities study; NOMAS, The Northern Manhattan Study; WHICAP, Washington Heights-Inwood Community Aging Project; EDIS, Epidemiology of Dementia In Singapore; MCS, Memory Clinic in Singapore; FHS, Framingham Heart Study.

We identified 14 variants at one locus associated with global brain arterial diameter at genome-wide significance (P<5×10^−8^; Figure 1A). This locus mapped to an intron of *CNNM2* (Cyclin and CBS Domain Divalent Metal Cation Transport Mediator 2). One copy of the C allele (minor allele frequency [MAF], 0.42) for the lead single-nucleotide polymorphism (SNP) rs7921574 was associated with 6% increased global brain arterial diameter (P=1.54×10^−8^) (Table 2; Figure S1A). We also identified 2 intronic variants in *TCF25* (Transcription Factor 25) associated with posterior brain arterial diameter at genome-wide significance (Figure 1B). One copy of the C allele (MAF= 0.29) for the lead SNP rs35994878 was associated with 11% increased posterior brain arterial diameter (P=2.94×10^−8^) (Table 2; Figure S1B). We did not observe any genome-wide significant association for anterior brain arterial diameter (Figure 1C). No genomic inflation was observed for any of the brain arterial diameter analyses (Figure S2). We also performed ancestry-specific regional Manhattan plots for top SNPs in global (rs7921574), anterior (rs7921574), and posterior (rs35994878) diameter meta-analysis, respectively (Figure S3, S4, and S5), but did not observe ancestry-specific genome-wide significant associations.

**Table 2.** Variants (P <5×10^−8^) associated with brain arterial diameter

| SNP | Chr | Position (hg 19) | Nearest gene | Relation to gene | Allele1 | Allele2 | AF | beta | se | P value | CADD |
| --- | --- | --- | --- | --- | --- | --- | --- | --- | --- | --- | --- |
| **Global** |  |  |  |  |  |  |  |  |  |  |  |
| rs7921574 | 10 | 104840970 | CNNM2 | ncRNA_intronic | C | T | 0.42 | 0.06 | 0.01 | 1.54E-08 | 0.33 |
| rs943035 | 10 | 104839152 | CNNM2 | ncRNA_intronic | C | T | 0.42 | 0.06 | 0.01 | 1.57E-08 | 0.71 |
| rs10883826* | 10 | 104830819 | CNNM2 | ncRNA_intronic | G | A | 0.42 | 0.06 | 0.01 | 1.58E-08 | 2.25 |
| rs943036 | 10 | 104836047 | CNNM2 | ncRNA_intronic | C | T | 0.42 | 0.06 | 0.01 | 1.66E-08 | 5.85 |
| rs10883824 | 10 | 104812897 | CNNM2 | ncRNA_intronic | G | A | 0.42 | 0.06 | 0.01 | 2.12E-08 | 2.98 |
| rs8139 | 10 | 104848123 | CNNM2 | ncRNA_intronic | A | G | 0.42 | 0.06 | 0.01 | 2.31E-08 | 7.82 |
| rs3740387 | 10 | 104849468 | CNNM2 | ncRNA_intronic | A | G | 0.42 | 0.06 | 0.01 | 2.33E-08 | 8.71 |
| rs10883817 | 10 | 104755431 | CNNM2 | ncRNA_intronic | A | G | 0.42 | 0.06 | 0.01 | 2.82E-08 | 0.37 |
| rs3902934 | 10 | 104746649 | CNNM2 | ncRNA_intronic | G | A | 0.42 | 0.06 | 0.01 | 2.84E-08 | 1.24 |
| rs10883823 | 10 | 104812331 | CNNM2 | ncRNA_intronic | C | T | 0.42 | 0.06 | 0.01 | 3.35E-08 | 0.25 |
| rs7911789 | 10 | 104756374 | CNNM2 | ncRNA_intronic | C | T | 0.42 | 0.06 | 0.01 | 3.84E-08 | 0.58 |
| rs67908413 | 10 | 104764989 | CNNM2 | ncRNA_intronic | C | T | 0.42 | 0.06 | 0.01 | 3.84E-08 | 6.84 |
| rs1890184 | 10 | 104748459 | CNNM2 | ncRNA_intronic | C | A | 0.42 | 0.06 | 0.01 | 4.26E-08 | 0.62 |
| rs10786733 | 10 | 104794947 | CNNM2 | ncRNA_intronic | A | G | 0.42 | 0.06 | 0.01 | 4.84E-08 | 4.21 |
| **Posterior** |  |  |  |  |  |  |  |  |  |  |  |
| rs35994878 | 16 | 89949033 | TCF25 | Intronic | C | T | 0.29 | 0.11 | 0.02 | 2.94E-08 | 1.06 |
| rs8061025 | 16 | 89948397 | TCF25 | Intronic | A | G | 0.29 | 0.11 | 0.02 | 4.49E-08 | 0.42 |

\* Independent variant associated with brain arterial diameter. Nearest gene with a functional protein or RNA product that either overlaps with the variant or for intergenic variants, the nearest genes up- and downstream, respectively. The statistics are based on Allele1. Allele1 indicates effect allele, allele2 is another allele. AF: allele 1 frequency. CADD, combined annotation dependent depletion score. Chr, chromosome.

**Figure 1.**
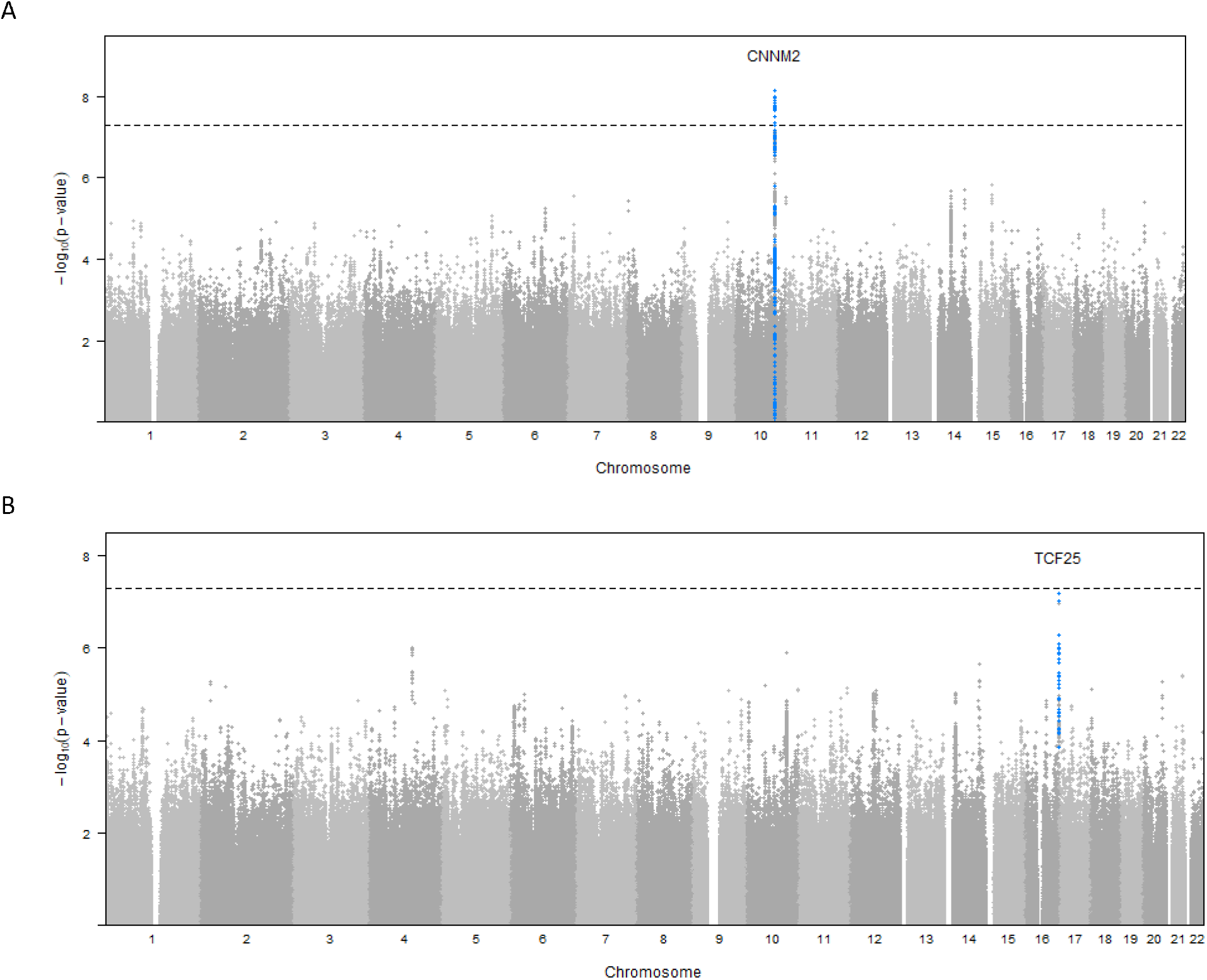
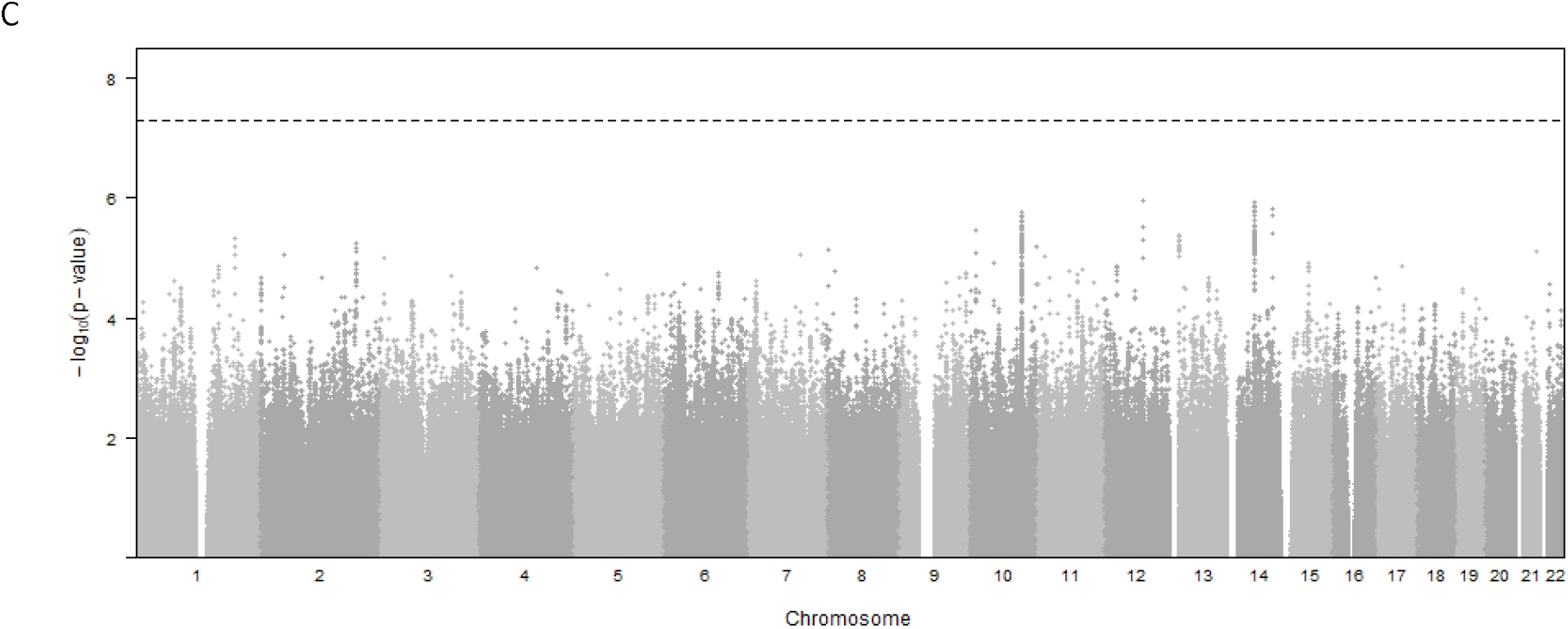
Genome-wide associations in brain arterial diameter. Manhattan plots for brain arterial diameter show combined genome-wide associations from 6 population-based studies.

In the trans-ancestry genome-wide association analysis for global brain arterial diameter, in addition to SNPs in *CNNM2*, we identified one genome-wide significant SNP near *NT5C2* (rs10748839; P=2.54×10^−8^) and one in *AS3MT* (rs10786721; P=4.97×10^−8^). All were located within 10q24.32. For anterior brain arterial diameter, we identified one locus at *ADAP1* (rs34217249; P=3.11×10^−8^) (Table 3). In the Hispanic-specific analysis, we identified a genome-wide significant locus at *LOC107986223* for global brain arterial diameter; three loci at *TGFBR2, LOC105374506*, and *LOC105376292* for posterior brain arterial diameter (Table S5). We did not observe genome-wide significant association in European, African, or Asian ancestries (Table S3, S4 and S6) for global, anterior, or posterior brain arterial diameter. No genomic inflation was observed in trans-ancestry analysis (Figure S6, S7, and S8).

**Table 3.** Trans-ancestry Genome-Wide Significant Associations

| SNP | Chr | Position (hg 19) | Nearest gene | Relation to gene | Allele1 | Allele2 | AF | P value | P_Het ANCS | P_Res Het | CADD |
| --- | --- | --- | --- | --- | --- | --- | --- | --- | --- | --- | --- |
| **Global** |  |  |  |  |  |  |  |  |  |  |  |
| rs10883805 | 10 | 104708251 | CNNM2 | ncRNA_intronic | C | T | 0.43 | 1.88E-08 | 0.003 | 0.333 | 4.39 |
| rs10748839 | 10 | 104953547 | NT5C2 | upstream | C | T | 0.42 | 2.54E-08 | 0.001 | 0.526 | 3.34 |
| rs3902934 | 10 | 104746649 | CNNM2 | ncRNA_intronic | G | A | 0.42 | 4.55E-08 | 0.026 | 0.329 | 1.24 |
| rs10883814 | 10 | 104737404 | CNNM2 | ncRNA_intronic | C | T | 0.43 | 4.79E-08 | 0.004 | 0.324 | 3.01 |
| rs12569617 | 10 | 104729996 | CNNM2 | ncRNA_intronic | C | T | 0.43 | 4.85E-08 | 0.004 | 0.323 | 2.87 |
| rs10786721 | 10 | 104654383 | AS3MT | ncRNA_intronic | A | C | 0.43 | 4.97E-08 | 0.004 | 0.384 | 4.52 |
| **Anterior** |  |  |  |  |  |  |  |  |  |  |  |
| rs34217249 | 7 | 960642 | ADAP1 | Intronic | A | G | 0.20 | 3.11E-08 | 2.03E-08 | 0.36 | 2.26 |

Nearest gene with a functional protein or RNA product that either overlaps with the variant or for intergenic variants, the nearest genes up- and downstream, respectively. The statistics are based on Allele1. Allele1 indicates effect allele, allele2 is another allele. AF: allele 1 frequency. CADD, combined annotation dependent depletion score. Chr, chromosome. P\_Het ANCS P-value for heterogeneity correlated with ancestry. P\_Res Het is the residual heterogeneity.

### Variant Effects Predictions on Protein Coding Sequence

We investigated the predicted deleterious effects of brain arterial diameter-associated loci using the Combined Annotation Dependent Depletion (CADD) scores. The SNPs and their proxies with CADD scores are shown in Table 2 and Table 3. We did not observe significant CADD score among the genome-wide significant loci associated with brain arterial diameter. A SNP associated with posterior brain arterial diameter in the *RAD52* region (rs140934041) showed significant CADD score (13.45) in Hispanic-specific analysis (Table S5). Additionally, SNPs associated with anterior brain arterial diameter in the *RAPGEF4* and posterior brain arterial diameter in *PODXL* region showed significant CADD scores (rs2290378, 16.31; rs888608, 16.63) in Asian-specific analysis (Table S6).

### Gene-Based Association Test and Gene-Set Enrichment

The Multi-Marker Analysis of GenoMic Annotation gene-based association analysis identified one locus associated with global brain arterial diameter (P<1.50×10^−5^) (Table S7). The significant associations for global brain arterial diameter included the GWAS located at *AS3MT, CNNM2, NT5C2, ARL3, TMEM180, C10orf32* and *C10orf32-ASMT*. Genes mapped to GWAS associations with P<1×10^−5^ were further investigated for gene-set enrichment (Table S8). Three genome-wide significant loci for global brain arterial diameter, *AS3MT, CNNM2* and *NT5C2*, were enriched in the white matter lesion progression gene set from GWAS catalog database (adjusted P=7.60×10^−7^).

### Tissue-Specific Colocalization Analyses

We performed colocalization analysis for the locus identified in the GWAS and MTAG analysis with gene expression using Genotype-Tissue Expression v8 eQTL data (Table S9). We identified SNPs associated with *AS3MT* and *C10orf32* expression and global brain arterial diameter in all 13 brain tissues. We also identified SNPs at TMEM180 in caudate basal ganglia, cerebellar hemisphere, nucleus accumbent basal ganglia, putamen basal ganglia, and spinal cord cervical c-1; SNPs at *CNNM2* in caudate basal ganglia tissues and SNPs at *NT5C2* and *ARL3* in cerebellum tissue, which colocalized with global brain arterial diameter. We also performed a transcriptome-wide association analysis for the loci identified in the GWAS with gene expression using BrainMeta project data in global, anterior, and posterior brain arterial diameter (Table 4). At the transcriptome-wide significance level (P<8.4×10^−6^), we identified SNPs associated with CNNM2 (P=6.17×10^−7^) and *AL356608*.*1* (P=6.6×10^−6^) expression in global brain arterial diameter (Figure 2A). We did not observe transcriptome-wide significant association in anterior or posterior brain arterial diameter (Figure 2B, 2C).

**Table 4.** Top 10 Co-localization of brain arterial diameter GWAS and eQTL associations

| Probe ID | Probe Chr | Gene | Probe_bp | SNP | SNP chr | SNP bp | Allele1 | Allele2 | AF | beta | se | p |
| --- | --- | --- | --- | --- | --- | --- | --- | --- | --- | --- | --- | --- |
| **Global** |  |  |  |  |  |  |  |  |  |  |  |  |
| ENSG00000148842.18 | 10 | CNNM2 | 104764015 | rs7897654 | 10 | 104662458 | C | T | 0.28 | 0.12 | 0.02 | 6.17E-07* |
| ENSG00000272912.1 | 10 | AL356608.1 | 104674752 | rs10786719 | 10 | 104637992 | G | A | 0.41 | -0.17 | 0.04 | 6.60E-06* |
| ENSG00000137760.15 | 11 | ALKBH8 | 107404960 | rs2037827 | 11 | 107408592 | C | T | 0.21 | -0.11 | 0.03 | 3.22E-04 |
| ENSG00000186715.11 | 1 | MST1L | 17089068 | rs7513616 | 1 | 17298496 | G | A | 0.43 | -0.06 | 0.02 | 6.88E-04 |
| ENSG00000140265.12 | 15 | ZSCAN29 | 43656796 | rs523156 | 15 | 43811843 | G | C | 0.53 | 0.08 | 0.03 | 7.75E-04 |
| ENSG00000014123.10 | 6 | UFL1 | 96986312 | rs11153023 | 6 | 96968525 | T | C | 0.15 | -0.12 | 0.03 | 8.19E-04 |
| ENSG00000237624.1 | 1 | OXCT2P1 | 39981395 | rs12028034 | 1 | 40039707 | A | G | 0.24 | 0.08 | 0.02 | 8.29E-04 |
| ENSG00000159363.19 | 1 | ATP13A2 | 17325438 | rs7513616 | 1 | 17298496 | G | A | 0.43 | 0.09 | 0.03 | 8.53E-04 |
| ENSG00000065717.15 | 19 | TLE2 | 3022635 | rs11150 | 19 | 2997897 | A | G | 0.17 | -0.17 | 0.05 | 9.46E-04 |
| ENSG00000268869.6 | 1 | ESPNP | 17030243 | rs7513616 | 1 | 17298496 | G | A | 0.43 | -0.12 | 0.04 | 1.36E-03 |
| **Anterior** |  |  |  |  |  |  |  |  |  |  |  |  |
| ENSG00000272912.1 | 10 | AL356608.1 | 104674752 | rs10786719 | 10 | 104637992 | G | A | 0.41 | -0.17 | 0.04 | 4.98E-05 |
| ENSG00000148842.18 | 10 | CNNM2 | 104764015 | rs7897654 | 10 | 104662458 | C | T | 0.28 | 0.10 | 0.02 | 6.62E-05 |
| ENSG00000162669.16 | 1 | HFM1 | 91798368 | rs17131417 | 1 | 91848784 | T | C | 0.11 | 0.06 | 0.02 | 7.78E-05 |
| ENSG00000162461.8 | 1 | SLC25A34 | 16065320 | rs41393951 | 1 | 16053493 | A | G | 0.31 | -0.05 | 0.01 | 3.75E-04 |
| ENSG00000137760.15 | 11 | ALKBH8 | 107404960 | rs2037827 | 11 | 107408592 | C | T | 0.21 | -0.11 | 0.03 | 3.79E-04 |
| ENSG00000065060.17 | 6 | UHRF1BP1 | 34805354 | rs6906129 | 6 | 34801160 | C | T | 0.51 | -0.05 | 0.02 | 4.60E-04 |
| ENSG00000156052.11 | 9 | GNAQ | 80488870 | rs4582625 | 9 | 80520544 | C | T | 0.26 | -0.08 | 0.02 | 6.44E-04 |
| ENSG00000257354.2 | 12 | AC048341.1 | 63006262 | rs17731893 | 12 | 63013773 | A | G | 0.18 | 0.27 | 0.08 | 6.70E-04 |
| ENSG00000231305.4 | 3 | AC112484.1 | 128584923 | rs789217 | 3 | 128593201 | A | G | 0.26 | 0.04 | 0.01 | 7.64E-04 |
| ENSG00000016402.13 | 6 | IL20RA | 137343712 | rs9494644 | 6 | 137403294 | G | C | 0.31 | 0.21 | 0.06 | 7.80E-04 |
| **Posterior** |  |  |  |  |  |  |  |  |  |  |  |  |
| ENSG00000138111.14 | 10 | MFSD13A | 104228977 | rs11593583 | 10 | 104228149 | G | A | 0.56 | -0.16 | 0.04 | 1.23E-04 |
| ENSG00000168386.18 | 3 | FILIP1L | 99691171 | rs6809988 | 3 | 99656615 | A | G | 0.22 | 0.13 | 0.03 | 2.75E-04 |
| ENSG00000213903.9 | 14 | LTB4R | 24783949 | rs11158632 | 14 | 24769663 | G | T | 0.22 | -0.23 | 0.06 | 4.01E-04 |
| ENSG00000148842.18 | 10 | CNNM2 | 104764015 | rs7897654 | 10 | 104662458 | C | T | 0.28 | 0.11 | 0.03 | 4.11E-04 |
| ENSG00000223959.8 | 16 | AFG3L1P | 90053782 | rs2270459 | 16 | 89979851 | A | C | 0.11 | -0.21 | 0.06 | 4.48E-04 |
| ENSG00000111615.14 | 12 | KRR1 | 75895030 | rs2070162 | 12 | 75900588 | G | A | 0.25 | -0.06 | 0.02 | 4.60E-04 |
| ENSG00000214043.8 | 12 | LINC02347 | 126940360 | rs17577161 | 12 | 126842742 | G | A | 0.34 | 0.06 | 0.02 | 5.97E-04 |
| ENSG00000255595.5 | 12 | AC007368.1 | 126797235 | rs17577161 | 12 | 126842742 | G | A | 0.34 | -0.07 | 0.02 | 6.18E-04 |
| ENSG00000258839.4 | 16 | MC1R | 89982954 | rs2270459 | 16 | 89979851 | A | C | 0.11 | 0.22 | 0.06 | 6.36E-04 |
| ENSG00000256310.1 | 12 | NDUFA5P6 | 127008983 | rs17577161 | 12 | 126842742 | G | A | 0.34 | 0.09 | 0.02 | 6.78E-04 |

Allele1 indicates effect allele, allele2 is another allele. AF: allele 1 frequency. \*Significant functional genes at P value < 8.4E-06

**Figure 2.**
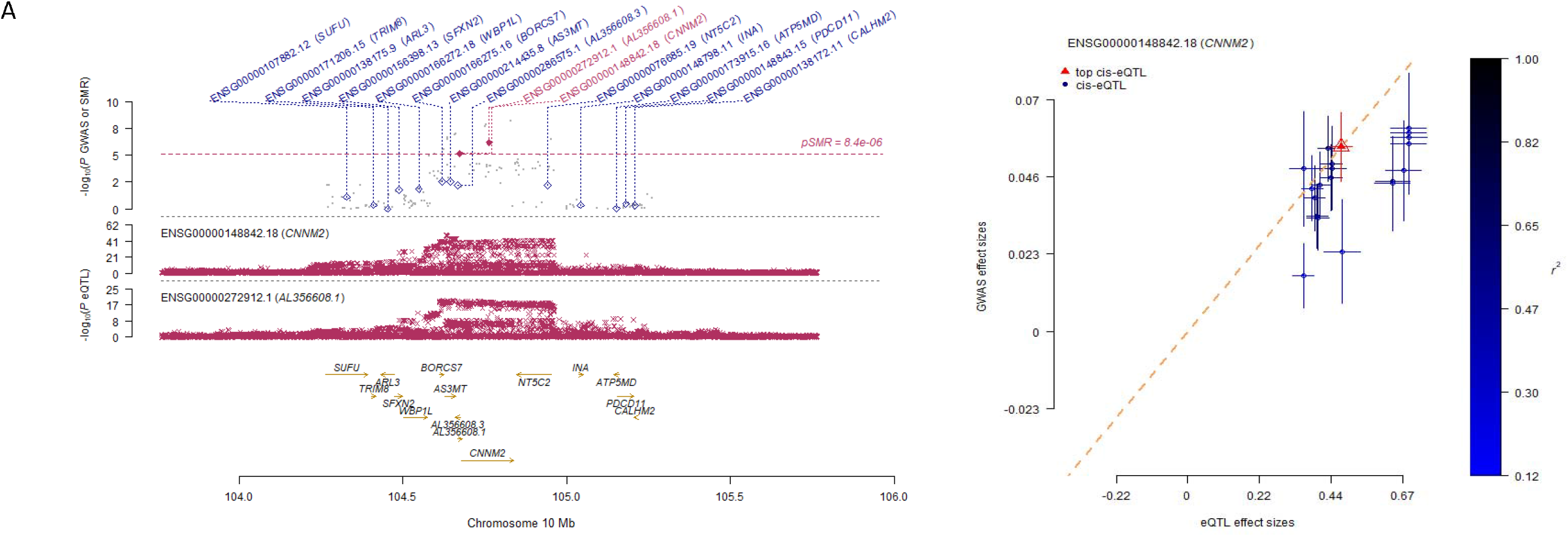
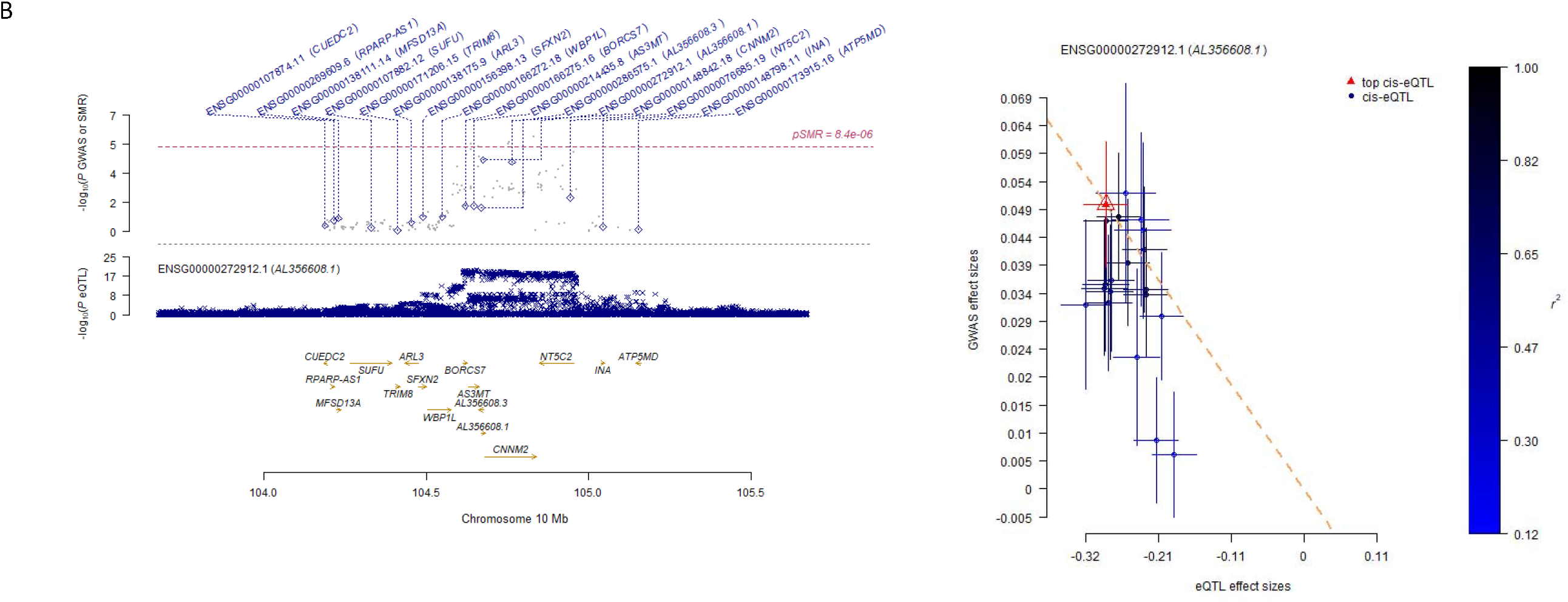
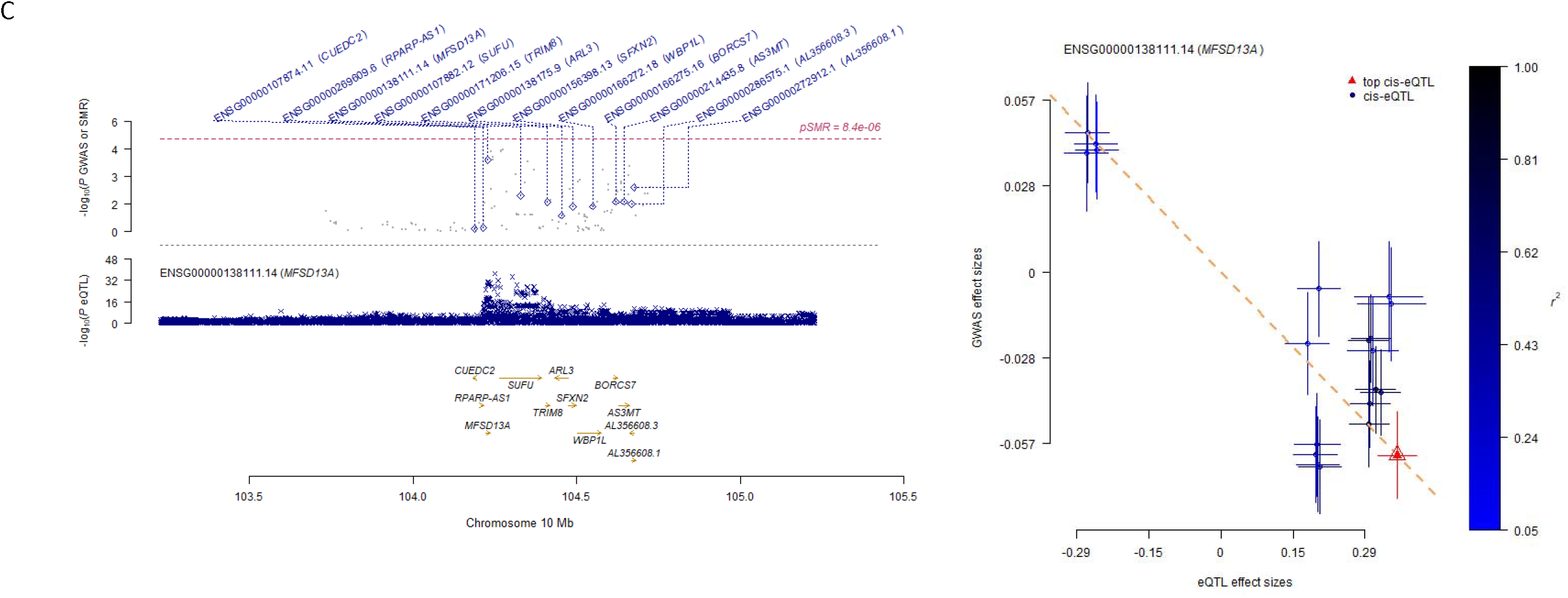
Locus plot and effect sizes plot of genome-wide association studies (GWAS) and expression quantitative trait locus (eQTL) associations.

### Causal Pathway from Brain Arterial Diameter to Alzheimer’s disease, stroke, and white matter hyperintensities volume

To establish a causal pathway from brain arterial diameter to Alzheimer’s disease, stroke, and white matter hyperintensities volume, we performed a Mendelian Randomization (MR) analysis (Table S10, S11). We did not observe any association of brain arterial diameter with Alzheimer’s disease, stroke, or white matter hyperintensities volume.

### Pleiotropic Locus for Anterior and Posterior Brain Arterial Diameter

MTAG analysis used the fixed-effect meta-analysis estimates for anterior and posterior brain arterial diameter. Since global brain arterial diameter is the average of anterior and posterior brain arterial diameter, the global estimate was excluded from multivariate analysis. No genomic inflation was observed in trans-ancestry analysis (Figure S9). MTAG results of joint analysis brain arterial diameter did not show any genome-wide significant SNPs (Table S12, S13).

## Discussion

This is the first study to examine the genetic determinants of brain arterial diameter in an ancestrally diverse population, where we identified associations of novel genetic loci with brain arterial diameter genetic architecture. Beyond mapping to the nearest genes, we also showed the biological impact of our findings using in silico functional analyses. Our results demonstrated that multiple genetic loci were coupled with gene expression information, which imply biologically relevant pathways.

We identified a novel brain arterial diameter locus at 10q24.32 mapped to *CNNM2. CNNM2* encodes Cyclin M2, which is a member of magnesium (Mg^2+^) transporters. As an abundant intracellular divalent cation in the human body, magnesium (Mg^2+^) plays an important role in numerous biological processes such as the synthesis of RNA, DNA and protein, and the production and storage of cellular energy ^42^. *CNNM2* is involved in brain development, neurological functioning and Mg^2+^ homeostasis ^43^. Heterozygous variants in the *CNNM2* gene can cause renal hypomagnesemia (HOMG6 [MIM 613882]), seizures, and intellectual disability (HOMGSMR1 [MIM 616418]) ^44^. In our study, variant rs7897654 was associated with decreased brain arterial diameter (β=-0.06) and an increased *CNNM2* expression (β=0.02) (Table 4). The variant rs7897654 colocalized with eQTLS of *CNNM2*, confirming its functional relationship to this gene. Our study also identified a variant in *TCF25* associated at genome-wide significance with posterior brain arterial diameter. *TCF25* is a member of the basic helix-loop-helix (bHLH) family of transcription factors that are important in embryonic development ^45^. These two results suggest that the effects of these genetic variants on arterial size might be present early in life, but how aging interacts with these variants remains unknown.

The *NT5C2* encodes a phosphatase involved in cellular purine metabolism, which is associated with disorders characterized by psychiatric and psychomotor disturbances ^46,47^. *NT5C2* has a high affinity for adenosine monophosphate and is involved in the extensive transcriptional programming which regulates cell maintenance, proliferation, migration, and differentiation during neurodevelopment ^48-51^. *NT5C2* has also been shown to negatively regulate phosphorylation of the alpha subunit of 5’-adenosine monophosphate-activated protein kinase (AMPK alpha) and protein translation^52^. Studies in the Chinese Han population report that *NT5C2* rs2148198 is associated with coronary heart disease susceptibility, and *NT5C2* rs11191580 is associated with schizophrenia and symptom severity ^53,54^. In addition, a zebrafish study provides evidence that *NT5C2* and *CNNM2* are most likely the causal genes within a blood pressure locus at the 10q24.32 ^55^. Our trans-ancestry GWAS analysis identified a significant variant rs10748839, mapped on the 2KB upstream of *NT5C2*, which is promoter variant that control expression of *NT5C2* ^68^.

The *AS3MT* gene, located in 10q24.32, encodes a cytosolic protein which is a cysteine rich enzyme that transfers a methyl group from S-adenosyl-L-methionine to trivalent arsenical ^56,57^. *AS3MT* plays an important role in catalysis of biomethylation of arsenic in vivo and in vitro. *AS3MT* is mainly expressed in human adrenal glands, liver, heart, kidney, and brain ^58^. Additionally, *AS3MT* expression is highly expressed in adult human neurons and astrocytes during human stem cell differentiation toward neuronal fates and in brains of patients with schizophrenia compared with controls ^59^ and with attention deficit or hyperactivity disorder^60^. Notably, *AS3MT* rs7085104 as a schizophrenia-associated risk SNP altered striatal dopamine synthesis capacity Moreover, the *AS3MT-CNNM2-NT5C2* gene cluster region is involved in etiology and pathogenesis of schizophrenia and the three genes have been confirmed as schizophrenia susceptibility gene cluster ^61,62^. Our study identified *AS3MT* rs10786721 variants with genome-wide significance in global brain arterial diameter. In addition, *AS3MT* rs72841270 is a lead variant associated with global brain arterial diameter in Hispanic-specific population. Whether neuronal connectivity or network formation indirectly or directly impacts brain arterial diameters is unclear but should be further studied.

Lysosomes play a critical role in maintenance of the integrity of neuronal function, and mutations in genes that contribute to lysosome formation, transport, and activity are associated with neurodegenerative disorders ^63,64^. Recently, the multi-subunit complex, BLOC-one-related complex (BORC), has been shown to be involved in positioning lysosomes within the cytoplasm, although the consequences of altered BORC function in adult animals have not been established ^65,66^. A study in mice identifies *BORCS7* (*C10orf32*) as a central factor in axonal transport of lysosomes and a possible target for improving disease-related disturbances in this important function; additionally, the Q87X mutation in the *BORCS7* subunit results in motor deficits and dystrophic axonopathy in mice ^67^. In our gene-based MAGMA analysis, the significant associations for global brain arterial diameter included *AS3MT, C10orf32, CNNM2* and *NT5C2*; we suspect that this four-gene cluster region may be involved with the etiology and pathogenesis of brain arterial diameter, but the underlying mechanism is not clear.

Our study is the first to explore the risk variants of brain arterial diameter in a large multi-ancestry GWAS. We detected novel SNPs located in genomic region 10q24.32 which are associated with brain arterial diameter. Due to the modest sample sizes of African, Asian, and Hispanic participants, the statistical power to detect ancestry-specific associations or functional associations in these ancestries were limited. Similarly, disentangling the effects of these variants on overall brain health versus AD specific pathways is difficult without functional analyses of genes related to arterial diameters, but exploring such pathways may reveal novel vascular contribution to Alzheimer’s disease and related dementias. Based on our results, we anticipate that the association between these genetic variants and brain arterial diameters will be consistent across populations, although the effect size might vary given the presence of common confounders such as vascular risk factors and environmental exposures.

## Conclusions

In summary, we identified a novel genome-wide significant locus for brain arterial diameter, *CNNM2, NT5C2* and *AS3MT*, in a large multi-ancestry population. Our study provides a potential biological mechanism for the association between 10q24.32 variation and brain arterial diameter. Identifying genes associated with these loci and their function may help us to elucidate the mechanism by which brain arterial diameters may influence cerebrovascular health.

## Methods

### Sampled populations

#### Atherosclerosis Risk in Communities (ARIC) study

The ARIC study is a population-based prospective cohort study of vascular risks and includes 15,792 persons aged 45-64 years at baseline (1987-89), randomly chosen from four US communities.^15^ Cohort members completed seven clinic examinations, conducted between 1987 and 2019. Written informed consent was provided by all study participants, and the study design and methods were approved by institutional review boards at the collaborating medical institutions (The Johns Hopkins University, Wake Forest University, University of Mississippi Medical Center, and University of Minnesota). Dementia and dementia subtypes were adjudicated beginning in 2011 using in-person interviews and cognitive testing, chart reviews and telephone surveys.^16^

#### The Northern Manhattan Study (NOMAS)

The NOMAS is an ongoing prospective cohort initially focused on determining the incidence of stroke and vascular events in a diverse urban population. Participants were recruited using random digit dialing between 1993 and 2001 with the following eligibility criteria: (1) age 40 or older, (2) clinically stroke free, and (3) resident of Northern Manhattan for at least 3 months. In person cognitive testing has been done three times since 2011 in surviving participants, Dementia was adjudicated by consensus between a neurologist and a neuropsychologist. The institutional review boards at Columbia University Medical Center and the University of Miami approved the study. All participants provided written informed consent.

#### Washington Heights–Inwood Columbia Aging Project (WHICAP study)

WHICAP is a prospective, population-based study of aging and dementia. Established through several recruitment waves, participants were first recruited in 1992 from a random sample of Medicare-eligible adults (age ≥65) residing in the neighborhoods of Washington Heights and Inwood in northern Manhattan. Participants are evaluated longitudinally every 18–24 months, with a comprehensive neuropsychological battery, medical and neurologic examination, and survey about health-related outcomes. ^17^ Dementia and dementia subtypes are adjudicated in a consensus conference that includes neurologists and neuropsychologists.

#### Epidemiology of Dementia In Singapore (EDIS study)

The EDIS study is a population-based cohort study conducted in southwestern Singapore between 2004 and 2011. It recruited participants who participated in the baseline visit of the Singapore Epidemiology of Eye Diseases (SEED) which comprised 10,033 adults of Chinese, Malay, and Indian ancestry, 40-80 years old ^18-21^. Briefly, the EDIS study consisted of three independent population cohorts with a common protocol. In all studies, individuals 40-80 years old were selected by an age-stratified random sampling method from a computer-generated random list of names provided by the Ministry of Home Affairs. The study was approved by the SERI Institutional Review Board. Written informed consent was obtained, in the preferred language of participants, by bilingual study coordinators prior to recruitment into the study.

#### Memory Clinic in Singapore (MCS study)

The MCS study included patients attending the National University Hospital (NUH) and St Luke’s Hospital memory clinics between 2009 and 2015. Patients were referred by primary care as well as secondary and tertiary care facilities because of consistent memory complaints and were assessed by a team of clinicians, psychologists, and nurses in the Memory Aging and Cognition Center, National University of Singapore.

#### Framingham Heart Study (FHS)

FHS started enrolling community-based participants in 1949. In 1971, all descendants of the original cohort (i.e., offspring cohort, requiring at least one parent from the original cohort) and their spouses were invited to participate in a follow-up study, and since then, they have been followed prospectively. The initial cohort consisted of 5124 men and women; 88% of survivors (3539/4031) participated in examination 7 in 1998-2001. Participants who survived to the 7th examination were invited to undergo a brain MRI (1999-2005), with a final sample of 2144 stroke-free, community-based participants. For these analyses, we used a FHS subsample with available MRA as part of the stroke case study.

### Measurement of brain arterial diameter

Brain magnetic resonance angiogram acquisition parameters by cohort are reported in supplemental Table 1. Brain arterial diameters and lengths were obtained from all available MRA images using commercial software (LAVA, Leiden University Medical Center, The Netherlands, build date Oct 19, 2018). Briefly, this software uses a flexible 3D tubular Non-Uniform Rational B-Splines model to automatically identify the margins of the arterial lumen based on voxel intensity ^22^ with excellent reliability.^23^ The 13 arterial segments measured included the bilateral intracranial internal carotid (ICA); middle cerebral (MCA), anterior cerebral (ACA), posterior cerebral (PCA), vertebral (VA), and posterior communicating (Pcomm) arteries, plus the basilar artery (BA). The location of measurement was aimed at the largest portion of a given segment free of focal stenosis, with good to excellent reliability. ^24^ For arteries visualized in the axial source MRA images but not large enough to be reconstructed, we systematically assigned the smallest measured diameters for the artery in the sample minus 10%. We counted arteries not visualized in axial source MRA images to create a score of absent arteries. We transformed each artery diameter distribution into normal scores and obtained the global (all 13 arteries), anterior (ICA, MCA, ACA and Pcomm if available) and posterior (VA, BA and PCA if available) arterial diameter scores by cohort as the principal dependent variable.

### Genotyping and Imputation

Detailed description of genotyping, quality control and imputation in each study is provided in supplemental Table 2. All analyses were conducted on autosomal chromosomes. Genotypes with missing rate greater than 10%, significant Hardy-Weinberg Equilibrium p-value (HWE p-value <5×10^−8^), or poor imputation quality (r^2^<0.3) were excluded from the analyses.

### Genome-wide Association Analysis

In each study and self-reported racial/ancestry strata, linear regression models were used to test the association between genetic variants and brain arterial diameter (global, anterior and posterior scores) using an additive genetic model, adjusted for sex, age, head size, number of absent arteries and population-specific principal components of ancestry (PCs). Genome-wide association studies (GWAS) results were subjected to quality control analyses using EasyQC (Winkler et al., 2013) and combined by meta-analysis using a fixed-effect inverse-variance-based method implemented in METAL ^25^ Variants with minor allele frequencies (MAF) < 1% and those do not present in at least two studies were excluded after the meta-analyses. Cross-study heterogeneity was assessed using Cochran’s Q-test, and variants with heterogeneity p-value <0.05 were excluded. A trans-ancestry meta-analysis of GWAS was conducted to account for heterogeneity in allelic effect that is correlated with ancestry by Meta-Regression of Multi-Ethnic Genetic Association (MR-MEGA). ^26^ Ancestry-specific meta-analyses were also performed to identify ancestry-specific variants. Multi-Trait Analysis of GWAS (MTAG) tool ^27^ was used for multivariate analysis of anterior and posterior brain arterial diameter to boost the statistical power to detect genetic associations. An association with a p-value < 5×10^−8^ was considered genome-wide significant, whereas p-value < 1.0×10^−5^ were used as suggestive evidence for marker associations.

### Gene-based Association Analysis and Gene-set Enrichment

We performed a gene-based association analysis based on summary statistics using Multi-marker Analysis of GenoMic Annotation (MAGMA v.1.07),^28^ implemented by FUnctional Mapping and Annotation (FUMA).^29^ Variants with p-value < 1×10^−5^ were mapped to the nearest gene within 50kb or an expression quantitative trait locus (eQTL) genes in Genotype-Tissue Expression (GTEx) project data version 8 (v8)^30^ from brain tissues: amygdala, anterior cingulate cortex (BA24), caudate (basal ganglia), cerebellar hemisphere, cerebellum, cortex, frontal Cortex (BA9), hippocampus, hypothalamus, nucleus accumbens (basal ganglia), putamen (basal ganglia), spinal cord (cervical c-1), substantia nigra. Mapped genes were then tested for tissue specificity in 30 general GTEx tissues using the pre-calculated differentially expressed gene (DEG) sets integrated in the GENE2FUNC of FUMA. ^29^ Hypergeometric enrichment tests were performed via GENE2FUNC against pre-defined gene sets obtained from Molecular signatures database (MsigDB) ^31^, WikiPathways ^32^ and GWAS catalog.^33^ We used Bonferroni corrected p-value < 0.05 to define statistical significance in gene-based analyses.

### Identification of Genomic Risk Loci and Deleteriousness of Lead SNPs

Variants that had at least suggestive evidence (p-value <10^−5^) were filtered and LD-clumped at r^2^<0.1 to identify independent loci using FUMA’s SNP2GENE function ^29^ based on the relevant 1000G reference. To investigate the protein coding consequences of lead independent variants associated with brain arterial diameter, the Combined Annotation Dependent Depletion (CADD) score was estimated. We used the threshold of 12.37 to determine whether a lead variant was deleterious. ^34^ When the CADD score of a lead variant was smaller than 12.37, we assessed whether its proxy variants (r^2^>0.8) were deleterious instead.

### Pleiotropic Association Analysis with Gene Expression

We used the SMR software ^35^ to test for pleiotropic association between brain arterial diameter traits and gene expression. We used summary-level data from our GWAS analyses and data on expression quantitative trait loci (eQTL) from the BrainMeta project version 2. ^36^ There are 5967 cis-eQTLs with eQTL□p-value<5×10^−8^. We used a Bonferroni corrected p-value < 8.4×10^−6^ (0.05/5967) to define statistical significance in pleiotropic association analyses.

### Two Sample Mendelian Randomization Analysis

We conducted a two-sample Mendelian randomization (MR) analysis using genetic instruments from the present analyses to assess whether brain arterial diameter is a causal factor for Alzheimer’s disease, stroke and white matter hyperintensities volume. The summary statistics for Alzheimer’s disease, stroke and white matter hyperintensities volume were used in this analysis. ^37-39^ To avoid bias driven by correlated instruments, variants with brain arterial diameter association p-value <1.0×10^−5^ were LD-clumped at r^2^ < 0.01 ^40^ against the 1000 Genome LD reference calculated for African, European, Asian and Hispanic populations. Variants with MAFs < 0.01 in the reference population were excluded from MR analysis. Causal association was primarily evaluated using the inverse-variance weighted method. To assess the presence of horizontal pleiotropy (i.e, that variants influence the outcome trait via independent pathways other than the exposure trait), we used the simple mode method, weighted mode method, inverse-variance weighted method (IVW), median-based method, and MR-Egger method. All MR analyses were performed using the “TwoSampleMR” R package. ^41^

## Data Availability

All data produced in the present study are available upon reasonable request to the authors.

## Declarations

### Ethics approval and consent to participate

The included studies have been approved by local ethics committees: Atherosclerosis Risk in Communities (ARIC) study: The institutional review board at The Johns Hopkins University, Wake Forest University, University of Mississippi Medical Center, and University of Minnesota; The Northern Manhattan Study (NOMAS): The institutional review boards at Columbia University Medical Center and the University of Miami; Washington Heights-Inwood Columbia Aging Project (WHICAP): The institutional review boards at Columbia University Medical Center; Epidemiology of Dementia In Singapore (EDIS): The institutional review boards at Singapore Eye Research Institute; Memory Clinic in Singapore (MCS): The institutional review boards at National University Hospital; Framingham Heart Study (FHS): The institutional review boards at Boston Medical Center.

### Consent for publication

Informed consent has been obtained from all participants included in the analyzed studies.

### Availability of data and materials

Studies participating in this meta-analysis have separate and specific data request and approval policies, depending on local, national, and international laws and regulations. Because of restrictions based on such privacy laws and regulations and informed consent of the participants, data cannot be made freely available in a public repository for any of the participating studies. Requests for information on procedures and formal data requests can be submitted to investigators from the corresponding author (Jose Gutierrez).

### Competing interests

The authors declare that they have no competing interests.

### Funding

This investigation was supported by National Institutes of Health grant R01 AG057709. The Atherosclerosis Risk in Communities (ARIC) Study is carried out as a collaborative study supported by National Heart, Lung, and Blood Institute contracts (75N92022D00001, 75N92022D00002, 75N92022D00003, 75N92022D00004, 75N92022D00005). The ARIC Neurocognitive Study is supported by U01HL096812, U01HL096814, U01HL096899, U01HL096902, and U01HL096917 from the NIH (NHLBI, NINDS, NIA and NIDCD). Funding was also supported by R01AG054491, R01HL087641 and R01HL086694; National Human Genome Research Institute contract U01HG004402; and National Institutes of Health contract HHSN268200625226C. Infrastructure was partly supported by Grant Number UL1RR025005, a component of the National Institutes of Health and NIH Roadmap for Medical Research. The Northern Manhattan Study (NOMAS) was supported by National Institutes of Health (R01 AG066162, R01 NS36286, R01 NS29993). The Washington Heights–Inwood Columbia Aging Project (WHICAP) was supported by National Institutes of Health (R01 AG072474, R01 AG037212, RF1 AG054023). The Epidemiology of Dementia In Singapore (EIDS) study was supported by the National Medical Research Council, Singapore (NMRC/CG/NUHS/2010 [Grant no: R-184-006-184-511]). The Memory Clinic in Singapore (MCS) was supported by U01 AG052409. The Framingham Heart Study (FHS) was supported by National Heart, Lung and Blood Institute contracts (N01 HC25195, HHSN268201500001I, 75N92019D00031) with additional support from National Institutes of Health grants (R01 AG047645, R01 HL131029) and an American Heart Association Award (15GPSGC24800006).

### Authors’ contributions

ML performed statistical analyses, and drafted and revised the manuscript. FK, SS, AS, YQ, ET, SS, JR participated in data acquisition and revised the manuscript. DS, HA, QY, AB, SH, TR, JJ, GT participated in data analysis and interpretation and revised the manuscript. AB, ME, RS, CC, BW, MF were responsible for obtaining funding and revising the manuscript. JG was responsible for the study concept and design, obtaining funding, and drafting and revising the manuscript. All authors read and approved the final manuscript.

## Acknowledgements

The authors thank the staff and participants of the ARIC, NOMAS, WHICAP, EDIS, MCS and FHS studies for their important contributions.

## Notes

### Competing Interest Statement

The authors have declared no competing interest.

### Summary of Updates

Author name updated

