## Supplementary material for "Chromosome 10q24.32 Variants Associate with Brain Arterial Diameters in Diverse Populations: A Genome-Wide Association Study": supp_tables_MRA_meta_12072022.docx

**Supplemental Material**

| **Supplemental Tables** | |
| --- | --- |
| Supplemental Table 1 | Genotype method |
| Supplemental Table 2 | MRA Specifications Brain Arterial Diameter Measurement |
| Supplemental Table 3 | Lead variants from European-specific meta-analysis |
| Supplemental Table 4 | Lead variants from African-specific meta-analysis |
| Supplemental Table 5 | Lead variants from Hispanic-specific meta-analysis |
| Supplemental Table 6 | Lead variants from Asian-specific meta-analysis |
| Supplemental Table 7 | Significant Associations from Gene-Based MAGMA Analysis |
| Supplemental Table 8 | Enriched Gene Sets for Global Brain Arterial Diameter |
| Supplemental Table 9 | Co-localization of Brain Arterial Diameter Loci with Tissue-Specific Gene Expression |
| Supplemental Table 10 | Mendelian Randomization Analysis between Global Brain Arterial Diameter and Alzheimer’s disease, Stroke, and White Matter Hyperintensities Volume |
| Supplemental Table 11 | Results for Five Mendelian Randomization Methods between Global Brain Arterial Diameter and Alzheimer's disease, Stroke, and White Matter Hyperintensities Volume |
| Supplemental Table 12 | Multi-Trait Analysis of Anterior Brain Arterial Diameter |
| Supplemental Table 13 | Multi-Trait Analysis of Posterior Brain Arterial Diameter |
| **Supplemental Figures** | |
| Supplemental Figure 1 | Forest plot and PM plot comparing brain arterial diameter estimate for multiple cohorts and meta-analysis estimate for SNP chr10:104840970 (rs7921574) and chr16:89949033 (rs35994878) |
| Supplemental Figure 2 | QQ plot and Manhattan plot of MR-MEGA all study meta-analysis |
| Supplemental Figure 3 | Locus zoom plot for top SNP rs7921574 in a global brain arterial diameter meta-analysis |
| Supplemental Figure 4 | Locus zoom plot for top SNP rs35994878 in posterior brain arterial diameter meta-analysis |
| Supplemental Figure 5 | Locus zoom plot for top SNP rs34217429 in anterior brain arterial diameter meta-analysis |
| Supplemental Figure 6 | QQ plot and Manhattan plot of global brain arterial diameter meta-analysis |
| Supplemental Figure 7 | QQ plot and Manhattan plot of anterior brain arterial diameter meta-analysis |
| Supplemental Figure 8 | QQ plot and Manhattan plot of posterior brain arterial diameter meta-analysis |
| Supplemental Figure 9 | QQ plot and Manhattan plot of for multivariate analysis of anterior and posterior brain arterial diameter |

**Supplemental tables and Supplemental Figures**

Supplemental table 1 Genotype method

| Cohort | Platform | Imputation reference | Ancestry | Number of post-QC SNPs included in the meta-analysis | Sample size |
| --- | --- | --- | --- | --- | --- |
| ARIC | Affymetrix 6.0 array | 1000 Genomes Phase 3 | European | 9,325,362 | 1163 |
|  |  |  | African | 14,165,061 | 402 |
| NOMAS | Affymetrix 6.0 array | 1000 Genomes Phase 3 | European | 6,034,689 | 151 |
|  |  |  | African | 9,379,418 | 181 |
|  |  |  | Hispanic | 12,074,793 | 760 |
| WHICAP | Illumina Omni Express | 1000 Genomes Phase 3 | European | 7,354,750 | 130 |
|  |  |  | Hispanic | 5,623,875 | 160 |
| EDIS | OmniExpress | 1000 Genomes Phase 3 | Asian/Chinese | 4,840,090 | 74 |
|  | Illumina Human610-Quad v1.0 |  | Asian/Malay | 6,379,450 | 231 |
|  |  |  | Asian/Indian | 6,879,279 | 211 |
|  |  |  | Asian/Chinese | 5,591,186 | 131 |
| MCS | Human610-Quad v1.0 | 1000 Genomes Phase 3 | Asian | 7,835,581 | 350 |
| FHS | Affymetrix GeneChip Human Mapping 500K Array | Haplotype Reference Consortium | European | 7,032,734 | 206 |

ARIC, Atherosclerosis Risk in Communities study; NOMAS, The Northern Manhattan Study; WHICAP, Washington Heights-Inwood Community Aging Project; EIDS, Epidemiology of Dementia In Singapore; MCS, Memory Clinic in Singapore; FHS, Framingham Heart Study.

Supplemental table 2 MRA Specifications Brain Arterial Diameter Measurement

| Cohort | MRI machine | Ac matrix | Flip angle | TR/TE | slice thick |
| --- | --- | --- | --- | --- | --- |
| ARIC | 3.0T Siemens | 320x320 | 18° | 21/3.7 | 0.5mm |
| NOMAS | 1.5T Philips | 256x228 | 25° | 20/2.7 | 1mm |
| WHICAP | 3.0T Philips | 520x247 | 20° | 25/3.5 | 0.8mm |
| EDIS | 3.0T Siemens | 320x320 | 18° | 21/3.7 | 0.5mm |
| MCS | 3.0T Siemens | 320x320 | 18° | 21/3.7 | 0.5mm |
| FHS | 1.5T Siemens | 192x256 | 12° | 21/3.7 | 1.5mm |

ARIC, Atherosclerosis Risk in Communities study; NOMAS, The Northern Manhattan Study; WHICAP, Washington Heights-Inwood Community Aging Project; EIDS, Epidemiology of Dementia In Singapore; MCS, Memory Clinic in Singapore; FHS, Framingham Heart Study.

Supplemental table 3 Lead variants from European-specific meta-analysis

| SNP | Chr | Position (hg 19) | Nearest gene | Relation to gene | Allele1 | Allele2 | AF | beta | se | P value | CADD |
| --- | --- | --- | --- | --- | --- | --- | --- | --- | --- | --- | --- |
| **Global** |  |  |  |  |  |  |  |  |  |  |  |
| rs10133360 | 14 | 34120460 | NPAS3 | ncRNA_intronic | C | T | 0.483 | 0.09 | 0.02 | 8.74E-08 | 0.84 |
| rs2075857 | 11 | 1250221 | MUC5B | ncRNA_intronic | A | G | 0.128 | 0.14 | 0.03 | 3.08E-07 | 2.31 |
| rs2075858 | 11 | 1250242 | MUC5B | ncRNA_intronic | T | G | 0.126 | 0.14 | 0.03 | 3.30E-07 | 1.84 |
| **Anterior** |  |  |  |  |  |  |  |  |  |  |  |
| rs281857 | 10 | 12963972 | CCDC3 | intronic | C | G | 0.072 | 0.18 | 0.03 | 1.73E-07 | 0.95 |
| **Posterior** |  |  |  |  |  |  |  |  |  |  |  |
| rs2075857 | 11 | 1250221 | MUC5B | intronic | A | G | 0.129 | 0.19 | 0.04 | 3.50E-07 | 2.3 |
| rs2075858 | 11 | 1250242 | MUC5B | intronic | T | G | 0.127 | 0.19 | 0.04 | 4.05E-07 | 1.84 |

Nearest gene with a functional protein or RNA product that either overlaps with the variant or for intergenic variants, the nearest genes up- and downstream, respectively. The statistics are based on Allele1. Allele1 indicates effect allele, allele2 is another allele. AF: allele 1 frequency. CADD, combined annotation dependent depletion score. Chr, chromosome.

Supplemental table 4 Lead variants from African-specific meta-analysis

| SNP | Chr | Position (hg 19) | Nearest gene | Relation to gene | Allele1 | Allele2 | AF | beta | se | P value | CADD |
| --- | --- | --- | --- | --- | --- | --- | --- | --- | --- | --- | --- |
| **Global** |  |  |  |  |  |  |  |  |  |  |  |
| rs478704 | 11 | 78473376 | TENM4 | intronic | T | C | 0.169 | 0.17 | 0.03 | 1.55E-07 | 6.52 |
| rs34074386 | 7 | 25945254 | LOC105375199 | intergenic | T | G | 0.128 | 0.18 | 0.04 | 2.38E-07 | 0.51 |
| **Anterior** |  |  |  |  |  |  |  |  |  |  |  |
| rs6945955 | 7 | 27370919 | HNRNPA1P73 | upstream | T | C | 0.099 | 0.23 | 0.04 | 1.14E-07 | 0.64 |
| rs10498548 | 14 | 81126016 | CEP128 | intronic | A | G | 0.726 | 0.16 | 0.03 | 2.07E-07 | 2.61 |
| rs17342675 | 16 | 56103090 | LOC105371281 | intronic | A | T | 0.074 | 0.29 | 0.06 | 3.29E-07 | 4.81 |
| rs12407465 | 1 | 61083444 | LINC01748 | intronic | C | T | 0.879 | 0.22 | 0.04 | 4.69E-07 | 0.63 |
| **Posterior** |  |  |  |  |  |  |  |  |  |  |  |
| rs7478716 | 11 | 69195256 | LOC105369370 | intergenic | T | C | 0.227 | 0.24 | 0.04 | 6.74E-08 | 1.14 |
| rs6771025 | 3 | 16587477 | LINC00690 | downstream | T | C | 0.923 | 0.35 | 0.07 | 3.12E-07 | 4.63 |
| rs72975853 | 3 | 139446261 | NMNAT3 | intergenic | C | T | 0.939 | 0.35 | 0.07 | 3.68E-07 | 4.62 |
| rs9911562 | 17 | 8288381 | RPL26 | intronic | C | T | 0.847 | 0.26 | 0.05 | 4.53E-07 | 5.68 |

Nearest gene with a functional protein or RNA product that either overlaps with the variant or for intergenic variants, the nearest genes up- and downstream, respectively. The statistics are based on Allele1. Allele1 indicates effect allele, allele2 is another allele. AF: allele 1 frequency. CADD, combined annotation dependent depletion score. Chr, chromosome.

Supplemental table 5 Lead variants from Hispanic-specific meta-analysis

| SNP | Chr | Position (hg 19) | Nearest gene | Relation to gene | Allele1 | Allele2 | AF | beta | se | P value | CADD |
| --- | --- | --- | --- | --- | --- | --- | --- | --- | --- | --- | --- |
| **Global** |  |  |  |  |  |  |  |  |  |  |  |
| rs12503381 | 4 | 32435842 | LOC107986223 | intergenic | G | T | 0.071 | 0.21 | 0.04 | 9.99E-09 | 4.95 |
| rs72841270 | 10 | 104642237 | AS3MT | ncRNA_intronic | G | T | 0.177 | 0.13 | 0.02 | 4.28E-07 | 9.15 |
| **Anterior** |  |  |  |  |  |  |  |  |  |  |  |
| rs72691989 | 9 | 4555774 | SLC1A1 | intronic | T | C | 0.029 | 0.41 | 0.08 | 1.15E-07 | 1.69 |
| rs17756784 | 9 | 4555611 | SLC1A1 | intronic | A | T | 0.029 | 0.41 | 0.08 | 1.17E-07 | 10.44 |
| rs17756747 | 9 | 4554871 | SLC1A1 | intronic | A | T | 0.029 | 0.41 | 0.08 | 1.20E-07 | 7.85 |
| rs113788025 | 9 | 4553576 | SPATA6L | intergenic | G | A | 0.029 | 0.4 | 0.08 | 1.30E-07 | 2.54 |
| rs12503381 | 4 | 32435842 | LOC107986223 | intergenic | G | T | 0.029 | 0.21 | 0.04 | 4.29E-07 | 4.95 |
| **Posterior** |  |  |  |  |  |  |  |  |  |  |  |
| rs58692095 | 3 | 30552484 | TGFBR2 | Intergenic | T | C | 0.927 | 0.47 | 0.08 | 3.55E-09 | 4.04 |
| rs7625124 | 3 | 30552811 | TGFBR2 | Intergenic | C | T | 0.929 | 0.46 | 0.08 | 4.93E-09 | 2.62 |
| rs79148902 | 3 | 30550535 | TGFBR2 | Intergenic | T | A | 0.929 | 0.45 | 0.08 | 1.25E-08 | 0.11 |
| rs72847492 | 3 | 30550516 | TGFBR2 | Intergenic | G | A | 0.929 | 0.45 | 0.08 | 1.27E-08 | 0.10 |
| rs75945022 | 3 | 30553795 | TGFBR2 | Intergenic | A | T | 0.928 | 0.45 | 0.08 | 1.28E-08 | 3.99 |
| rs57865541 | 2 | 41792322 | LOC105374506 | intronic | C | T | 0.948 | 0.49 | 0.09 | 2.73E-08 | 0.18 |
| rs114263903 | 9 | 132327176 | LOC105376292 | Intergenic | G | C | 0.966 | 0.66 | 0.12 | 4.20E-08 | 0.81 |
| rs72650728 | 12 | 993684 | WNK1 | intronic | A | G | 0.951 | 0.53 | 0.1 | 1.37E-07 | 0.05 |
| rs138861130 | 12 | 1029178 | RAD52 | intronic | A | G | 0.95 | 0.52 | 0.1 | 1.56E-07 | 3.55 |
| rs140934041 | 12 | 1075141 | RAD52 | intronic | T | C | 0.945 | 0.5 | 0.1 | 1.59E-07 | 13.45 |
| rs79183605 | 22 | 51157531 | SHANK3 | intronic | G | A | 0.915 | 0.41 | 0.08 | 1.88E-07 | 2.32 |
| rs117281960 | 12 | 945599 | WNK1 | intronic | T | A | 0.95 | 0.52 | 0.1 | 2.64E-07 | 0.02 |
| rs111353779 | 1 | 46814029 | NSUN4 | intronic | G | A | 0.944 | 0.41 | 0.08 | 3.40E-07 | 5.62 |
| rs7559991 | 2 | 218233291 | DIRC3 | intronic | T | C | 0.674 | 0.2 | 0.04 | 4.28E-07 | 7.01 |

Nearest gene with a functional protein or RNA product that either overlaps with the variant or for intergenic variants, the nearest genes up- and downstream, respectively. The statistics are based on Allele1. Allele1 indicates effect allele, allele2 is another allele. AF: allele 1 frequency. CADD, combined annotation dependent depletion score. Chr, chromosome.

Supplemental table 6 Lead variants from Asian-specific meta-analysis

| SNP | Chr | Position (hg 19) | Nearest gene | Relation to gene | Allele1 | Allele2 | AF | beta | se | P value | CADD |
| --- | --- | --- | --- | --- | --- | --- | --- | --- | --- | --- | --- |
| **Anterior** |  |  |  |  |  |  |  |  |  |  |  |
| rs78996903 | 2 | 173913032 | RAPGEF4 | intronic | T | A | 0.478 | 0.15 | 0.03 | 3.70E-07 | 5.64 |
| rs2290378 | 2 | 173913258 | RAPGEF4 | intronic | A | G | 0.478 | 0.15 | 0.03 | 3.70E-07 | 16.31 |
| **Posterior** |  |  |  |  |  |  |  |  |  |  |  |
| rs10257021 | 7 | 131320998 | PODXL | Intergenic | G | C | 0.778 | 0.16 | 0.03 | 7.18E-08 | 1.27 |
| rs10272608 | 7 | 131320880 | PODXL | Intergenic | C | T | 0.778 | 0.16 | 0.03 | 8.04E-08 | 2.69 |
| rs10272488 | 7 | 131320832 | PODXL | Intergenic | C | T | 0.780 | 0.16 | 0.03 | 8.44E-08 | 0.21 |
| rs10257102 | 7 | 131320871 | PODXL | Intergenic | A | G | 0.771 | 0.16 | 0.03 | 1.29E-07 | 0.07 |
| rs888607 | 7 | 131322409 | PODXL | intergenic | T | A | 0.774 | 0.15 | 0.03 | 2.86E-07 | 1.21 |
| rs888608 | 7 | 131323108 | PODXL | Intergenic | C | A | 0.775 | 0.15 | 0.03 | 3.52E-07 | 16.63 |
| rs1000202 | 7 | 131323797 | PODXL | Intergenic | C | T | 0.774 | 0.15 | 0.03 | 4.51E-07 | 0.42 |

Nearest gene with a functional protein or RNA product that either overlaps with the variant or for intergenic variants, the nearest genes up- and downstream, respectively. The statistics are based on Allele1. Allele1 indicates effect allele, allele2 is another allele. AF: allele 1 frequency. CADD, combined annotation dependent depletion score. Chr, chromosome.

Supplemental table 7 Significant Associations from Gene-Based MAGMA Analysis

| Ensembl gene ID | Gene Symbol | Chr | Start (hg 19) | End (hg 19) | NSNPS | p-value |
| --- | --- | --- | --- | --- | --- | --- |
| **Global** |  |  |  |  |  |  |
| ENSG00000214435 | AS3MT | 10 | 104629273 | 104661656 | 6 | 1.78E-23 |
| ENSG00000166275 | C10orf32 | 10 | 104613980 | 104624718 | 6 | 1.18E-15 |
| ENSG00000148842 | CNNM2 | 10 | 104678050 | 104849978 | 3 | 1.03E-08 |
| ENSG00000076685 | NT5C2 | 10 | 104845940 | 104953056 | 4 | 1.03E-08 |
| ENSG00000270316 | C10orf32-ASMT | 10 | 104614029 | 104661656 | 1 | 1.68E-07 |
| ENSG00000138111 | TMEM180 | 10 | 104221149 | 104236802 | 6 | 6.16E-06 |
| ENSG00000138175 | ARL3 | 10 | 104433488 | 104474164 | 6 | 1.51E-05 |
| MAGMA: Multi-marker Analysis of GenoMic Annotation. | | | | | | |

Supplemental table 8 Enriched Gene Sets for Global Brain Arterial Diameter

| Database | GeneSet | N | n | p-value | adjusted p | genes |
| --- | --- | --- | --- | --- | --- | --- |
| Positional gene sets (MsigDB c1) | chr10q24 | 119 | 7 | 3E-16 | 8.97E-14 | TMEM180, ARL3, C10orf32, C10orf32-ASMT, AS3MT, CNNM2, NT5C2 |
| GWAS catalog | Autism spectrum disorder or schizophrenia | 474 | 7 | 5.47E-12 | 7.60E-09 | TMEM180, ARL3, C10orf32, C10orf32-ASMT, AS3MT, CNNM2, NT5C2 |
| GWAS catalog | Waist-to-hip ratio adjusted for BMI (age >50) | 200 | 6 | 8.38E-12 | 7.60E-09 | TMEM180, ARL3, C10orf32, AS3MT, CNNM2, NT5C2 |
| GWAS catalog | Prostate cancer | 302 | 6 | 1.01E-10 | 6.13E-08 | TMEM180, ARL3, C10orf32, C10orf32-ASMT, AS3MT, CNNM2 |
| GWAS catalog | Waist-to-hip ratio adjusted for BMI | 346 | 6 | 2.30E-10 | 1.05E-07 | TMEM180, ARL3, C10orf32, AS3MT, CNNM2, NT5C2 |
| GWAS catalog | Mean arterial pressure | 131 | 5 | 2.89E-10 | 1.05E-07 | ARL3, C10orf32, AS3MT, CNNM2, NT5C2 |
| GWAS catalog | White matter lesion progression | 9 | 3 | 2.51E-09 | 7.60E-07 | AS3MT, CNNM2, NT5C2 |
| GWAS catalog | Schizophrenia | 704 | 6 | 1.64E-08 | 4.26E-06 | ARL3, C10orf32, C10orf32-ASMT, AS3MT, CNNM2, NT5C2 |
| GWAS catalog | Microalbuminuria | 28 | 3 | 9.77E-08 | 1.98E-05 | C10orf32, C10orf32-ASMT, CNNM2 |
| GWAS catalog | Coronary artery disease | 418 | 5 | 9.82E-08 | 1.98E-05 | C10orf32, C10orf32-ASMT, AS3MT, CNNM2, NT5C2 |
| GWAS catalog | Autism spectrum disorder, attention deficit-hyperactivity disorder, bipolar disorder, major depressive disorder, and schizophrenia (combined) | 45 | 3 | 4.22E-07 | 7.66E-05 | AS3MT, CNNM2, NT5C2 |
| GWAS catalog | Blood pressure | 90 | 3 | 3.47E-06 | 5.73E-04 | AS3MT, CNNM2, NT5C2 |
| GWAS catalog | Hypertension | 95 | 3 | 4.09E-06 | 6.18E-04 | C10orf32, CNNM2, NT5C2 |
| GWAS catalog | Cognitive ability, years of educational attainment or schizophrenia (pleiotropy) | 165 | 3 | 2.15E-05 | 3.00E-03 | C10orf32, C10orf32-ASMT, AS3MT |
| GWAS catalog | Coronary artery disease or large artery stroke | 22 | 2 | 2.64E-05 | 3.42E-03 | CNNM2, NT5C2 |
| GWAS catalog | Pulse pressure | 647 | 4 | 4.17E-05 | 5.05E-03 | ARL3, C10orf32, CNNM2, NT5C2 |
| GWAS catalog | Response to cognitive-behavioural therapy in major depressive disorder | 35 | 2 | 6.78E-05 | 7.46E-03 | CNNM2, NT5C2 |
| GWAS catalog | Systolic blood pressure | 738 | 4 | 6.99E-05 | 7.46E-03 | ARL3, AS3MT, CNNM2, NT5C2 |
| GWAS catalog | Creatine kinase levels | 46 | 2 | 1.18E-04 | 1.12E-02 | C10orf32-ASMT, CNNM2 |
| GWAS catalog | Smoking initiation | 46 | 2 | 1.18E-04 | 1.12E-02 | C10orf32, CNNM2 |
| GWAS catalog | Myocardial infarction | 55 | 2 | 1.69E-04 | 1.53E-02 | CNNM2, NT5C2 |
| GWAS catalog | Waist-to-hip ratio adjusted for BMI x sex x age interaction (4df test) | 341 | 3 | 1.86E-04 | 1.61E-02 | TMEM180, ARL3, AS3MT |
| GWAS catalog | Coronary heart disease | 70 | 2 | 2.74E-04 | 2.26E-02 | CNNM2, NT5C2 |
| GWAS catalog | Risk-taking tendency (4-domain principal component model) | 89 | 2 | 4.42E-04 | 3.49E-02 | TMEM180, AS3MT |
| GWAS catalog | Coronary artery disease (myocardial infarction, percutaneous transluminal coronary angioplasty, coronary artery bypass grafting, angina or chromic ischemic heart disease) | 94 | 2 | 4.93E-04 | 3.73E-02 | CNNM2, NT5C2 |
| GWAS catalog | Immature fraction of reticulocytes | 102 | 2 | 5.80E-04 | 4.21E-02 | C10orf32, C10orf32-ASMT |

N: Number of genes in the gene set, n: number of genes enriched

Supplemental table 9 Co-localization of Brain Arterial Diameter Loci with Tissue-Specific Gene Expression

| symbol | tissue | Risking Allele | chr | pos | p | FDR |
| --- | --- | --- | --- | --- | --- | --- |
| AS3MT | Brain_Amygdala | A | 10 | 104612335 | 2.36E-08 | 4.50E-06 |
| AS3MT | Brain_Amygdala | G | 10 | 104628873 | 2.36E-08 | 4.50E-06 |
| AS3MT | Brain_Amygdala | G | 10 | 104618695 | 8.00E-08 | 4.50E-06 |
| AS3MT | Brain_Amygdala | G | 10 | 104621068 | 8.00E-08 | 4.50E-06 |
| AS3MT | Brain_Amygdala | G | 10 | 104625237 | 8.00E-08 | 4.50E-06 |
| AS3MT | Brain_Amygdala | A | 10 | 104625886 | 8.00E-08 | 4.50E-06 |
| AS3MT | Brain_Amygdala | T | 10 | 104595719 | 1.04E-07 | 4.50E-06 |
| AS3MT | Brain_Amygdala | T | 10 | 104596396 | 1.04E-07 | 4.50E-06 |
| AS3MT | Brain_Amygdala | A | 10 | 104605892 | 1.04E-07 | 4.50E-06 |
| AS3MT | Brain_Amygdala | A | 10 | 104599323 | 2.43E-07 | 4.50E-06 |
| AS3MT | Brain_Anterior_cingulate_cortex_BA24 | A | 10 | 104612335 | 1.44E-15 | 2.77E-11 |
| AS3MT | Brain_Anterior_cingulate_cortex_BA24 | G | 10 | 104628873 | 1.44E-15 | 2.77E-11 |
| AS3MT | Brain_Anterior_cingulate_cortex_BA24 | G | 10 | 104618695 | 4.10E-14 | 2.77E-11 |
| AS3MT | Brain_Anterior_cingulate_cortex_BA24 | G | 10 | 104621068 | 4.10E-14 | 2.77E-11 |
| AS3MT | Brain_Anterior_cingulate_cortex_BA24 | G | 10 | 104625237 | 4.10E-14 | 2.77E-11 |
| AS3MT | Brain_Anterior_cingulate_cortex_BA24 | A | 10 | 104625886 | 4.10E-14 | 2.77E-11 |
| AS3MT | Brain_Anterior_cingulate_cortex_BA24 | T | 10 | 104595719 | 1.00E-12 | 2.77E-11 |
| AS3MT | Brain_Anterior_cingulate_cortex_BA24 | T | 10 | 104596396 | 1.00E-12 | 2.77E-11 |
| AS3MT | Brain_Anterior_cingulate_cortex_BA24 | A | 10 | 104605892 | 1.00E-12 | 2.77E-11 |
| AS3MT | Brain_Anterior_cingulate_cortex_BA24 | A | 10 | 104596924 | 1.16E-11 | 2.77E-11 |
| AS3MT | Brain_Anterior_cingulate_cortex_BA24 | G | 10 | 104597152 | 1.16E-11 | 2.77E-11 |
| AS3MT | Brain_Caudate_basal_ganglia | A | 10 | 104612335 | 4.89E-12 | 1.01E-07 |
| AS3MT | Brain_Caudate_basal_ganglia | G | 10 | 104628873 | 4.89E-12 | 1.01E-07 |
| AS3MT | Brain_Caudate_basal_ganglia | G | 10 | 104618695 | 9.64E-12 | 1.01E-07 |
| AS3MT | Brain_Caudate_basal_ganglia | G | 10 | 104621068 | 9.64E-12 | 1.01E-07 |
| AS3MT | Brain_Caudate_basal_ganglia | G | 10 | 104625237 | 9.64E-12 | 1.01E-07 |
| AS3MT | Brain_Caudate_basal_ganglia | A | 10 | 104625886 | 9.64E-12 | 1.01E-07 |
| AS3MT | Brain_Caudate_basal_ganglia | A | 10 | 104599323 | 3.88E-11 | 1.01E-07 |
| AS3MT | Brain_Caudate_basal_ganglia | T | 10 | 104596396 | 3.93E-11 | 1.01E-07 |
| AS3MT | Brain_Caudate_basal_ganglia | A | 10 | 104605892 | 3.93E-11 | 1.01E-07 |
| AS3MT | Brain_Caudate_basal_ganglia | A | 10 | 104596924 | 7.42E-11 | 1.01E-07 |
| AS3MT | Brain_Caudate_basal_ganglia | G | 10 | 104597152 | 7.42E-11 | 1.01E-07 |
| AS3MT | Brain_Cerebellar_Hemisphere | G | 10 | 104618695 | 1.74E-30 | 9.71E-24 |
| AS3MT | Brain_Cerebellar_Hemisphere | G | 10 | 104621068 | 1.74E-30 | 9.71E-24 |
| AS3MT | Brain_Cerebellar_Hemisphere | G | 10 | 104625237 | 1.74E-30 | 9.71E-24 |
| AS3MT | Brain_Cerebellar_Hemisphere | A | 10 | 104625886 | 1.74E-30 | 9.71E-24 |
| AS3MT | Brain_Cerebellar_Hemisphere | A | 10 | 104612335 | 5.53E-29 | 9.71E-24 |
| AS3MT | Brain_Cerebellar_Hemisphere | G | 10 | 104628873 | 5.53E-29 | 9.71E-24 |
| AS3MT | Brain_Cerebellar_Hemisphere | A | 10 | 104596924 | 1.95E-24 | 9.71E-24 |
| AS3MT | Brain_Cerebellar_Hemisphere | G | 10 | 104597152 | 1.95E-24 | 9.71E-24 |
| AS3MT | Brain_Cerebellar_Hemisphere | C | 10 | 104602215 | 1.95E-24 | 9.71E-24 |
| AS3MT | Brain_Cerebellar_Hemisphere | G | 10 | 104603588 | 1.95E-24 | 9.71E-24 |
| AS3MT | Brain_Cerebellar_Hemisphere | A | 10 | 104618524 | 6.70E-22 | 9.71E-24 |
| AS3MT | Brain_Cerebellum | G | 10 | 104618695 | 3.44E-42 | 6.43E-34 |
| AS3MT | Brain_Cerebellum | G | 10 | 104621068 | 3.44E-42 | 6.43E-34 |
| AS3MT | Brain_Cerebellum | G | 10 | 104625237 | 3.68E-42 | 6.43E-34 |
| AS3MT | Brain_Cerebellum | A | 10 | 104625886 | 3.68E-42 | 6.43E-34 |
| AS3MT | Brain_Cerebellum | A | 10 | 104612335 | 3.54E-40 | 6.43E-34 |
| AS3MT | Brain_Cerebellum | G | 10 | 104628873 | 3.54E-40 | 6.43E-34 |
| AS3MT | Brain_Cerebellum | A | 10 | 104618524 | 3.61E-35 | 6.43E-34 |
| AS3MT | Brain_Cerebellum | G | 10 | 104619448 | 3.61E-35 | 6.43E-34 |
| AS3MT | Brain_Cerebellum | C | 10 | 104620735 | 3.61E-35 | 6.43E-34 |
| AS3MT | Brain_Cerebellum | A | 10 | 104794947 | 5.54E-23 | 6.43E-34 |
| AS3MT | Brain_Cerebellum | T | 10 | 104811699 | 5.54E-23 | 6.43E-34 |
| AS3MT | Brain_Cortex | G | 10 | 104618695 | 3.84E-23 | 2.88E-19 |
| AS3MT | Brain_Cortex | G | 10 | 104621068 | 3.84E-23 | 2.88E-19 |
| AS3MT | Brain_Cortex | G | 10 | 104625237 | 3.84E-23 | 2.88E-19 |
| AS3MT | Brain_Cortex | A | 10 | 104625886 | 3.84E-23 | 2.88E-19 |
| AS3MT | Brain_Cortex | A | 10 | 104612335 | 4.14E-23 | 2.88E-19 |
| AS3MT | Brain_Cortex | G | 10 | 104628873 | 4.14E-23 | 2.88E-19 |
| AS3MT | Brain_Cortex | T | 10 | 104866863 | 1.30E-18 | 2.88E-19 |
| AS3MT | Brain_Cortex | A | 10 | 104599323 | 1.46E-18 | 2.88E-19 |
| AS3MT | Brain_Cortex | A | 10 | 104596924 | 1.77E-18 | 2.88E-19 |
| AS3MT | Brain_Cortex | G | 10 | 104597152 | 1.77E-18 | 2.88E-19 |
| AS3MT | Brain_Cortex | C | 10 | 104602215 | 1.77E-18 | 2.88E-19 |
| AS3MT | Brain_Cortex | G | 10 | 104603588 | 1.77E-18 | 2.88E-19 |
| AS3MT | Brain_Frontal_Cortex_BA9 | A | 10 | 104612335 | 1.64E-15 | 9.03E-11 |
| AS3MT | Brain_Frontal_Cortex_BA9 | G | 10 | 104628873 | 1.64E-15 | 9.03E-11 |
| AS3MT | Brain_Frontal_Cortex_BA9 | G | 10 | 104618695 | 6.82E-15 | 9.03E-11 |
| AS3MT | Brain_Frontal_Cortex_BA9 | G | 10 | 104621068 | 6.82E-15 | 9.03E-11 |
| AS3MT | Brain_Frontal_Cortex_BA9 | G | 10 | 104625237 | 6.82E-15 | 9.03E-11 |
| AS3MT | Brain_Frontal_Cortex_BA9 | A | 10 | 104625886 | 6.82E-15 | 9.03E-11 |
| AS3MT | Brain_Frontal_Cortex_BA9 | T | 10 | 104595719 | 4.73E-13 | 9.03E-11 |
| AS3MT | Brain_Frontal_Cortex_BA9 | T | 10 | 104596396 | 1.16E-12 | 9.03E-11 |
| AS3MT | Brain_Frontal_Cortex_BA9 | A | 10 | 104605892 | 1.16E-12 | 9.03E-11 |
| AS3MT | Brain_Frontal_Cortex_BA9 | A | 10 | 104596924 | 3.16E-12 | 9.03E-11 |
| AS3MT | Brain_Frontal_Cortex_BA9 | G | 10 | 104597152 | 3.16E-12 | 9.03E-11 |
| AS3MT | Brain_Hippocampus | A | 10 | 104612335 | 9.14E-11 | 1.64E-08 |
| AS3MT | Brain_Hippocampus | G | 10 | 104628873 | 9.14E-11 | 1.64E-08 |
| AS3MT | Brain_Hippocampus | G | 10 | 104618695 | 8.35E-10 | 1.64E-08 |
| AS3MT | Brain_Hippocampus | G | 10 | 104621068 | 8.35E-10 | 1.64E-08 |
| AS3MT | Brain_Hippocampus | G | 10 | 104625237 | 8.35E-10 | 1.64E-08 |
| AS3MT | Brain_Hippocampus | A | 10 | 104625886 | 8.35E-10 | 1.64E-08 |
| AS3MT | Brain_Hippocampus | A | 10 | 104618524 | 3.56E-08 | 1.64E-08 |
| AS3MT | Brain_Hippocampus | G | 10 | 104619448 | 3.56E-08 | 1.64E-08 |
| AS3MT | Brain_Hippocampus | C | 10 | 104620735 | 3.56E-08 | 1.64E-08 |
| AS3MT | Brain_Hippocampus | A | 10 | 104624072 | 3.56E-08 | 1.64E-08 |
| AS3MT | Brain_Hippocampus | G | 10 | 104624475 | 3.56E-08 | 1.64E-08 |
| AS3MT | Brain_Hypothalamus | A | 10 | 104612335 | 6.63E-21 | 4.87E-15 |
| AS3MT | Brain_Hypothalamus | G | 10 | 104628873 | 6.63E-21 | 4.87E-15 |
| AS3MT | Brain_Hypothalamus | G | 10 | 104618695 | 8.20E-21 | 4.87E-15 |
| AS3MT | Brain_Hypothalamus | G | 10 | 104621068 | 8.20E-21 | 4.87E-15 |
| AS3MT | Brain_Hypothalamus | G | 10 | 104625237 | 8.20E-21 | 4.87E-15 |
| AS3MT | Brain_Hypothalamus | A | 10 | 104625886 | 8.20E-21 | 4.87E-15 |
| AS3MT | Brain_Hypothalamus | T | 10 | 104866863 | 1.25E-20 | 4.87E-15 |
| AS3MT | Brain_Hypothalamus | A | 10 | 104618524 | 1.90E-17 | 4.87E-15 |
| AS3MT | Brain_Hypothalamus | G | 10 | 104619448 | 1.90E-17 | 4.87E-15 |
| AS3MT | Brain_Hypothalamus | C | 10 | 104620735 | 1.90E-17 | 4.87E-15 |
| AS3MT | Brain_Hypothalamus | A | 10 | 104624072 | 1.90E-17 | 4.87E-15 |
| AS3MT | Brain_Nucleus_accumbens_basal_ganglia | T | 10 | 104596396 | 3.71E-11 | 9.93E-08 |
| AS3MT | Brain_Nucleus_accumbens_basal_ganglia | A | 10 | 104605892 | 3.71E-11 | 9.93E-08 |
| AS3MT | Brain_Nucleus_accumbens_basal_ganglia | A | 10 | 104612335 | 3.80E-11 | 9.93E-08 |
| AS3MT | Brain_Nucleus_accumbens_basal_ganglia | G | 10 | 104628873 | 3.80E-11 | 9.93E-08 |
| AS3MT | Brain_Nucleus_accumbens_basal_ganglia | A | 10 | 104596924 | 4.85E-11 | 9.93E-08 |
| AS3MT | Brain_Nucleus_accumbens_basal_ganglia | G | 10 | 104597152 | 4.85E-11 | 9.93E-08 |
| AS3MT | Brain_Nucleus_accumbens_basal_ganglia | C | 10 | 104602215 | 4.85E-11 | 9.93E-08 |
| AS3MT | Brain_Nucleus_accumbens_basal_ganglia | G | 10 | 104603588 | 4.85E-11 | 9.93E-08 |
| AS3MT | Brain_Nucleus_accumbens_basal_ganglia | C | 10 | 104609365 | 4.85E-11 | 9.93E-08 |
| AS3MT | Brain_Nucleus_accumbens_basal_ganglia | C | 10 | 104609466 | 4.85E-11 | 9.93E-08 |
| AS3MT | Brain_Nucleus_accumbens_basal_ganglia | G | 10 | 104618695 | 5.43E-11 | 9.93E-08 |
| AS3MT | Brain_Putamen_basal_ganglia | A | 10 | 104612335 | 6.58E-13 | 1.79E-08 |
| AS3MT | Brain_Putamen_basal_ganglia | G | 10 | 104628873 | 6.58E-13 | 1.79E-08 |
| AS3MT | Brain_Putamen_basal_ganglia | G | 10 | 104618695 | 8.39E-13 | 1.79E-08 |
| AS3MT | Brain_Putamen_basal_ganglia | G | 10 | 104621068 | 8.39E-13 | 1.79E-08 |
| AS3MT | Brain_Putamen_basal_ganglia | G | 10 | 104625237 | 8.39E-13 | 1.79E-08 |
| AS3MT | Brain_Putamen_basal_ganglia | A | 10 | 104625886 | 8.39E-13 | 1.79E-08 |
| AS3MT | Brain_Putamen_basal_ganglia | A | 10 | 104599323 | 1.33E-11 | 1.79E-08 |
| AS3MT | Brain_Putamen_basal_ganglia | T | 10 | 104596396 | 2.21E-11 | 1.79E-08 |
| AS3MT | Brain_Putamen_basal_ganglia | A | 10 | 104605892 | 2.21E-11 | 1.79E-08 |
| AS3MT | Brain_Putamen_basal_ganglia | A | 10 | 104596924 | 2.77E-11 | 1.79E-08 |
| AS3MT | Brain_Putamen_basal_ganglia | G | 10 | 104597152 | 2.77E-11 | 1.79E-08 |
| AS3MT | Brain_Spinal_cord_cervical_c-1 | A | 10 | 104612335 | 1.30E-16 | 1.89E-11 |
| AS3MT | Brain_Spinal_cord_cervical_c-1 | G | 10 | 104628873 | 1.30E-16 | 1.89E-11 |
| AS3MT | Brain_Spinal_cord_cervical_c-1 | G | 10 | 104625237 | 2.12E-16 | 1.89E-11 |
| AS3MT | Brain_Spinal_cord_cervical_c-1 | A | 10 | 104625886 | 2.12E-16 | 1.89E-11 |
| AS3MT | Brain_Spinal_cord_cervical_c-1 | G | 10 | 104618695 | 8.42E-16 | 1.89E-11 |
| AS3MT | Brain_Spinal_cord_cervical_c-1 | G | 10 | 104621068 | 8.42E-16 | 1.89E-11 |
| AS3MT | Brain_Spinal_cord_cervical_c-1 | T | 10 | 104595719 | 1.40E-13 | 1.89E-11 |
| AS3MT | Brain_Spinal_cord_cervical_c-1 | T | 10 | 104596396 | 1.40E-13 | 1.89E-11 |
| AS3MT | Brain_Spinal_cord_cervical_c-1 | A | 10 | 104605892 | 1.40E-13 | 1.89E-11 |
| AS3MT | Brain_Spinal_cord_cervical_c-1 | T | 10 | 104866863 | 2.80E-13 | 1.89E-11 |
| AS3MT | Brain_Spinal_cord_cervical_c-1 | A | 10 | 104599323 | 3.68E-13 | 1.89E-11 |
| AS3MT | Brain_Substantia_nigra | A | 10 | 104612335 | 9.95E-13 | 7.38E-08 |
| AS3MT | Brain_Substantia_nigra | G | 10 | 104628873 | 9.95E-13 | 7.38E-08 |
| AS3MT | Brain_Substantia_nigra | G | 10 | 104618695 | 4.21E-12 | 7.38E-08 |
| AS3MT | Brain_Substantia_nigra | G | 10 | 104621068 | 4.21E-12 | 7.38E-08 |
| AS3MT | Brain_Substantia_nigra | G | 10 | 104625237 | 4.21E-12 | 7.38E-08 |
| AS3MT | Brain_Substantia_nigra | A | 10 | 104625886 | 4.21E-12 | 7.38E-08 |
| AS3MT | Brain_Substantia_nigra | T | 10 | 104595719 | 5.99E-11 | 7.38E-08 |
| AS3MT | Brain_Substantia_nigra | T | 10 | 104596396 | 5.99E-11 | 7.38E-08 |
| AS3MT | Brain_Substantia_nigra | A | 10 | 104605892 | 5.99E-11 | 7.38E-08 |
| AS3MT | Brain_Substantia_nigra | A | 10 | 104596924 | 2.12E-10 | 7.38E-08 |
| AS3MT | Brain_Substantia_nigra | G | 10 | 104597152 | 2.12E-10 | 7.38E-08 |
| C10orf32 | Brain_Anterior_cingulate_cortex_BA24 | A | 10 | 104612335 | 1.89E-07 | 1.69E-07 |
| C10orf32 | Brain_Anterior_cingulate_cortex_BA24 | G | 10 | 104628873 | 1.89E-07 | 1.69E-07 |
| C10orf32 | Brain_Anterior_cingulate_cortex_BA24 | G | 10 | 104618695 | 2.71E-07 | 1.69E-07 |
| C10orf32 | Brain_Anterior_cingulate_cortex_BA24 | G | 10 | 104621068 | 2.71E-07 | 1.69E-07 |
| C10orf32 | Brain_Anterior_cingulate_cortex_BA24 | G | 10 | 104625237 | 2.71E-07 | 1.69E-07 |
| C10orf32 | Brain_Anterior_cingulate_cortex_BA24 | A | 10 | 104625886 | 2.71E-07 | 1.69E-07 |
| C10orf32 | Brain_Caudate_basal_ganglia | A | 10 | 104612335 | 2.89E-19 | 9.23E-22 |
| C10orf32 | Brain_Caudate_basal_ganglia | G | 10 | 104628873 | 2.89E-19 | 9.23E-22 |
| C10orf32 | Brain_Caudate_basal_ganglia | G | 10 | 104618695 | 7.43E-18 | 9.23E-22 |
| C10orf32 | Brain_Caudate_basal_ganglia | G | 10 | 104621068 | 7.43E-18 | 9.23E-22 |
| C10orf32 | Brain_Caudate_basal_ganglia | G | 10 | 104625237 | 7.43E-18 | 9.23E-22 |
| C10orf32 | Brain_Caudate_basal_ganglia | A | 10 | 104625886 | 7.43E-18 | 9.23E-22 |
| C10orf32 | Brain_Caudate_basal_ganglia | A | 10 | 104618524 | 2.36E-16 | 9.23E-22 |
| C10orf32 | Brain_Caudate_basal_ganglia | G | 10 | 104619448 | 2.36E-16 | 9.23E-22 |
| C10orf32 | Brain_Caudate_basal_ganglia | C | 10 | 104620735 | 2.36E-16 | 9.23E-22 |
| C10orf32 | Brain_Caudate_basal_ganglia | A | 10 | 104624072 | 2.36E-16 | 9.23E-22 |
| C10orf32 | Brain_Cerebellar_Hemisphere | A | 10 | 104612335 | 2.95E-16 | 1.36E-18 |
| C10orf32 | Brain_Cerebellar_Hemisphere | G | 10 | 104628873 | 2.95E-16 | 1.36E-18 |
| C10orf32 | Brain_Cerebellar_Hemisphere | G | 10 | 104618695 | 3.56E-16 | 1.36E-18 |
| C10orf32 | Brain_Cerebellar_Hemisphere | G | 10 | 104621068 | 3.56E-16 | 1.36E-18 |
| C10orf32 | Brain_Cerebellar_Hemisphere | G | 10 | 104625237 | 3.56E-16 | 1.36E-18 |
| C10orf32 | Brain_Cerebellar_Hemisphere | A | 10 | 104625886 | 3.56E-16 | 1.36E-18 |
| C10orf32 | Brain_Cerebellar_Hemisphere | T | 10 | 104596396 | 3.84E-14 | 1.36E-18 |
| C10orf32 | Brain_Cerebellar_Hemisphere | A | 10 | 104605892 | 3.84E-14 | 1.36E-18 |
| C10orf32 | Brain_Cerebellar_Hemisphere | A | 10 | 104596924 | 4.43E-14 | 1.36E-18 |
| C10orf32 | Brain_Cerebellar_Hemisphere | G | 10 | 104597152 | 4.43E-14 | 1.36E-18 |
| C10orf32 | Brain_Cerebellum | A | 10 | 104612335 | 3.32E-22 | 1.19E-21 |
| C10orf32 | Brain_Cerebellum | G | 10 | 104628873 | 3.32E-22 | 1.19E-21 |
| C10orf32 | Brain_Cerebellum | G | 10 | 104625237 | 8.69E-22 | 1.19E-21 |
| C10orf32 | Brain_Cerebellum | A | 10 | 104625886 | 8.69E-22 | 1.19E-21 |
| C10orf32 | Brain_Cerebellum | G | 10 | 104618695 | 3.07E-21 | 1.19E-21 |
| C10orf32 | Brain_Cerebellum | G | 10 | 104621068 | 3.07E-21 | 1.19E-21 |
| C10orf32 | Brain_Cerebellum | A | 10 | 104618524 | 1.60E-20 | 1.19E-21 |
| C10orf32 | Brain_Cerebellum | G | 10 | 104619448 | 1.60E-20 | 1.19E-21 |
| C10orf32 | Brain_Cerebellum | C | 10 | 104620735 | 1.60E-20 | 1.19E-21 |
| C10orf32 | Brain_Cerebellum | A | 10 | 104624072 | 1.60E-20 | 1.19E-21 |
| C10orf32 | Brain_Cortex | A | 10 | 104612335 | 2.00E-25 | 5.95E-26 |
| C10orf32 | Brain_Cortex | G | 10 | 104628873 | 2.00E-25 | 5.95E-26 |
| C10orf32 | Brain_Cortex | G | 10 | 104618695 | 2.67E-24 | 5.95E-26 |
| C10orf32 | Brain_Cortex | G | 10 | 104621068 | 2.67E-24 | 5.95E-26 |
| C10orf32 | Brain_Cortex | G | 10 | 104625237 | 2.67E-24 | 5.95E-26 |
| C10orf32 | Brain_Cortex | A | 10 | 104625886 | 2.67E-24 | 5.95E-26 |
| C10orf32 | Brain_Cortex | A | 10 | 104618524 | 1.97E-19 | 5.95E-26 |
| C10orf32 | Brain_Cortex | G | 10 | 104619448 | 1.97E-19 | 5.95E-26 |
| C10orf32 | Brain_Cortex | C | 10 | 104620735 | 1.97E-19 | 5.95E-26 |
| C10orf32 | Brain_Cortex | A | 10 | 104624072 | 1.97E-19 | 5.95E-26 |
| C10orf32 | Brain_Frontal_Cortex_BA9 | A | 10 | 104612335 | 3.33E-15 | 8.55E-21 |
| C10orf32 | Brain_Frontal_Cortex_BA9 | G | 10 | 104628873 | 3.33E-15 | 8.55E-21 |
| C10orf32 | Brain_Frontal_Cortex_BA9 | G | 10 | 104618695 | 3.94E-15 | 8.55E-21 |
| C10orf32 | Brain_Frontal_Cortex_BA9 | G | 10 | 104621068 | 3.94E-15 | 8.55E-21 |
| C10orf32 | Brain_Frontal_Cortex_BA9 | G | 10 | 104625237 | 3.94E-15 | 8.55E-21 |
| C10orf32 | Brain_Frontal_Cortex_BA9 | A | 10 | 104625886 | 3.94E-15 | 8.55E-21 |
| C10orf32 | Brain_Frontal_Cortex_BA9 | A | 10 | 104618524 | 1.29E-13 | 8.55E-21 |
| C10orf32 | Brain_Frontal_Cortex_BA9 | G | 10 | 104619448 | 1.29E-13 | 8.55E-21 |
| C10orf32 | Brain_Frontal_Cortex_BA9 | C | 10 | 104620735 | 1.29E-13 | 8.55E-21 |
| C10orf32 | Brain_Frontal_Cortex_BA9 | A | 10 | 104624072 | 1.29E-13 | 8.55E-21 |
| C10orf32 | Brain_Hippocampus | A | 10 | 104612335 | 1.12E-06 | 3.87E-06 |
| C10orf32 | Brain_Hippocampus | G | 10 | 104628873 | 1.12E-06 | 3.87E-06 |
| C10orf32 | Brain_Hippocampus | G | 10 | 104618695 | 2.88E-06 | 3.87E-06 |
| C10orf32 | Brain_Hippocampus | G | 10 | 104621068 | 2.88E-06 | 3.87E-06 |
| C10orf32 | Brain_Hippocampus | G | 10 | 104625237 | 2.88E-06 | 3.87E-06 |
| C10orf32 | Brain_Hippocampus | A | 10 | 104625886 | 2.88E-06 | 3.87E-06 |
| C10orf32 | Brain_Hippocampus | A | 10 | 104618524 | 4.94E-06 | 3.87E-06 |
| C10orf32 | Brain_Hippocampus | G | 10 | 104619448 | 4.94E-06 | 3.87E-06 |
| C10orf32 | Brain_Hippocampus | C | 10 | 104620735 | 4.94E-06 | 3.87E-06 |
| C10orf32 | Brain_Hippocampus | A | 10 | 104624072 | 4.94E-06 | 3.87E-06 |
| C10orf32 | Brain_Hypothalamus | A | 10 | 104612335 | 3.00E-10 | 1.12E-09 |
| C10orf32 | Brain_Hypothalamus | G | 10 | 104628873 | 3.00E-10 | 1.12E-09 |
| C10orf32 | Brain_Hypothalamus | A | 10 | 104618524 | 3.87E-10 | 1.12E-09 |
| C10orf32 | Brain_Hypothalamus | G | 10 | 104619448 | 3.87E-10 | 1.12E-09 |
| C10orf32 | Brain_Hypothalamus | C | 10 | 104620735 | 3.87E-10 | 1.12E-09 |
| C10orf32 | Brain_Hypothalamus | A | 10 | 104624072 | 3.87E-10 | 1.12E-09 |
| C10orf32 | Brain_Hypothalamus | G | 10 | 104624475 | 3.87E-10 | 1.12E-09 |
| C10orf32 | Brain_Hypothalamus | C | 10 | 104625178 | 3.87E-10 | 1.12E-09 |
| C10orf32 | Brain_Hypothalamus | G | 10 | 104629465 | 3.87E-10 | 1.12E-09 |
| C10orf32 | Brain_Hypothalamus | T | 10 | 104633114 | 3.87E-10 | 1.12E-09 |
| C10orf32 | Brain_Nucleus_accumbens_basal_ganglia | A | 10 | 104618524 | 9.51E-10 | 1.44E-10 |
| C10orf32 | Brain_Nucleus_accumbens_basal_ganglia | G | 10 | 104619448 | 9.51E-10 | 1.44E-10 |
| C10orf32 | Brain_Nucleus_accumbens_basal_ganglia | C | 10 | 104620735 | 9.51E-10 | 1.44E-10 |
| C10orf32 | Brain_Nucleus_accumbens_basal_ganglia | A | 10 | 104624072 | 9.51E-10 | 1.44E-10 |
| C10orf32 | Brain_Nucleus_accumbens_basal_ganglia | G | 10 | 104624475 | 9.51E-10 | 1.44E-10 |
| C10orf32 | Brain_Nucleus_accumbens_basal_ganglia | C | 10 | 104625178 | 9.51E-10 | 1.44E-10 |
| C10orf32 | Brain_Nucleus_accumbens_basal_ganglia | G | 10 | 104629465 | 9.51E-10 | 1.44E-10 |
| C10orf32 | Brain_Nucleus_accumbens_basal_ganglia | T | 10 | 104633114 | 9.51E-10 | 1.44E-10 |
| C10orf32 | Brain_Nucleus_accumbens_basal_ganglia | C | 10 | 104635344 | 9.51E-10 | 1.44E-10 |
| C10orf32 | Brain_Nucleus_accumbens_basal_ganglia | G | 10 | 104635595 | 9.51E-10 | 1.44E-10 |
| C10orf32 | Brain_Putamen_basal_ganglia | A | 10 | 104612335 | 8.91E-15 | 5.81E-19 |
| C10orf32 | Brain_Putamen_basal_ganglia | G | 10 | 104628873 | 8.91E-15 | 5.81E-19 |
| C10orf32 | Brain_Putamen_basal_ganglia | G | 10 | 104618695 | 9.80E-14 | 5.81E-19 |
| C10orf32 | Brain_Putamen_basal_ganglia | G | 10 | 104621068 | 9.80E-14 | 5.81E-19 |
| C10orf32 | Brain_Putamen_basal_ganglia | G | 10 | 104625237 | 9.80E-14 | 5.81E-19 |
| C10orf32 | Brain_Putamen_basal_ganglia | A | 10 | 104625886 | 9.80E-14 | 5.81E-19 |
| C10orf32 | Brain_Putamen_basal_ganglia | A | 10 | 104618524 | 1.82E-11 | 5.81E-19 |
| C10orf32 | Brain_Putamen_basal_ganglia | G | 10 | 104619448 | 1.82E-11 | 5.81E-19 |
| C10orf32 | Brain_Putamen_basal_ganglia | C | 10 | 104620735 | 1.82E-11 | 5.81E-19 |
| C10orf32 | Brain_Putamen_basal_ganglia | A | 10 | 104624072 | 1.82E-11 | 5.81E-19 |
| C10orf32 | Brain_Spinal_cord_cervical_c-1 | A | 10 | 104612335 | 8.18E-14 | 5.11E-12 |
| C10orf32 | Brain_Spinal_cord_cervical_c-1 | G | 10 | 104628873 | 8.18E-14 | 5.11E-12 |
| C10orf32 | Brain_Spinal_cord_cervical_c-1 | G | 10 | 104625237 | 1.00E-13 | 5.11E-12 |
| C10orf32 | Brain_Spinal_cord_cervical_c-1 | A | 10 | 104625886 | 1.00E-13 | 5.11E-12 |
| C10orf32 | Brain_Spinal_cord_cervical_c-1 | G | 10 | 104618695 | 1.26E-13 | 5.11E-12 |
| C10orf32 | Brain_Spinal_cord_cervical_c-1 | G | 10 | 104621068 | 1.26E-13 | 5.11E-12 |
| C10orf32 | Brain_Spinal_cord_cervical_c-1 | T | 10 | 104595719 | 5.03E-13 | 5.11E-12 |
| C10orf32 | Brain_Spinal_cord_cervical_c-1 | T | 10 | 104596396 | 5.03E-13 | 5.11E-12 |
| C10orf32 | Brain_Spinal_cord_cervical_c-1 | A | 10 | 104605892 | 5.03E-13 | 5.11E-12 |
| C10orf32 | Brain_Spinal_cord_cervical_c-1 | A | 10 | 104596924 | 8.74E-13 | 5.11E-12 |
| C10orf32 | Brain_Substantia_nigra | A | 10 | 104612335 | 7.53E-06 | 0.0001782 |
| C10orf32 | Brain_Substantia_nigra | G | 10 | 104628873 | 7.53E-06 | 0.0001782 |
| ARL3 | Brain_Cerebellum | A | 10 | 104612335 | 2.47E-07 | 3.19E-05 |
| ARL3 | Brain_Cerebellum | G | 10 | 104628873 | 2.47E-07 | 3.19E-05 |
| ARL3 | Brain_Cerebellum | G | 10 | 104625237 | 3.47E-07 | 3.19E-05 |
| ARL3 | Brain_Cerebellum | A | 10 | 104625886 | 3.47E-07 | 3.19E-05 |
| ARL3 | Brain_Cerebellum | G | 10 | 104618695 | 3.67E-07 | 3.19E-05 |
| ARL3 | Brain_Cerebellum | G | 10 | 104621068 | 3.67E-07 | 3.19E-05 |
| ARL3 | Brain_Cerebellum | T | 10 | 104596396 | 2.49E-06 | 3.19E-05 |
| ARL3 | Brain_Cerebellum | A | 10 | 104605892 | 2.49E-06 | 3.19E-05 |
| ARL3 | Brain_Cerebellum | G | 10 | 104658992 | 3.50E-06 | 3.19E-05 |
| ARL3 | Brain_Cerebellum | T | 10 | 104659018 | 3.50E-06 | 3.19E-05 |
| TMEM180 | Brain_Caudate_basal_ganglia | C | 10 | 104844872 | 1.02E-06 | 1.70E-08 |
| TMEM180 | Brain_Caudate_basal_ganglia | A | 10 | 104850632 | 1.17E-06 | 1.70E-08 |
| TMEM180 | Brain_Caudate_basal_ganglia | C | 10 | 104836047 | 6.16E-06 | 1.70E-08 |
| TMEM180 | Brain_Caudate_basal_ganglia | C | 10 | 104839152 | 6.16E-06 | 1.70E-08 |
| TMEM180 | Brain_Caudate_basal_ganglia | A | 10 | 104848123 | 6.16E-06 | 1.70E-08 |
| TMEM180 | Brain_Caudate_basal_ganglia | A | 10 | 104849468 | 6.16E-06 | 1.70E-08 |
| TMEM180 | Brain_Caudate_basal_ganglia | C | 10 | 104855656 | 6.16E-06 | 1.70E-08 |
| TMEM180 | Brain_Caudate_basal_ganglia | G | 10 | 104866958 | 6.16E-06 | 1.70E-08 |
| TMEM180 | Brain_Caudate_basal_ganglia | G | 10 | 104871361 | 6.16E-06 | 1.70E-08 |
| TMEM180 | Brain_Caudate_basal_ganglia | T | 10 | 104872547 | 6.16E-06 | 1.70E-08 |
| CNNM2 | Brain_Caudate_basal_ganglia | T | 10 | 104647849 | 8.66E-05 | 0.0079426 |
| CNNM2 | Brain_Caudate_basal_ganglia | C | 10 | 104649729 | 8.66E-05 | 0.0079426 |
| CNNM2 | Brain_Caudate_basal_ganglia | A | 10 | 104652495 | 8.66E-05 | 0.0079426 |
| NT5C2 | Brain_Cerebellum | T | 10 | 104866863 | 9.49E-05 | 0.0013007 |

Supplemental table 10 Mendelian Randomization Analysis between Global Brain Arterial Diameter and Alzheimer’s disease, Stroke, and White Matter Hyperintensities Volume

| Outcome | Heterogeneity Test | | |  | Pleiotropy Test | | |
| --- | --- | --- | --- | --- | --- | --- | --- |
|  | Q | df | P |  | egger intercept | se | p |
| Alzheimer's disease | 6.881 | 14 | 0.939 |  | 6.62E-05 | 8.07E-05 | 0.426 |
| Stroke | 5.211 | 14 | 0.988 |  | -9.29E-05 | 0.0002 | 0.731 |
| White matter hyperintensities volume | 10.07 | 14 | 0.757 |  | 0.012 | 0.007 | 0.082 |

Supplemental table 11 Results for Five Mendelian Randomization Methods between Global Brain Arterial Diameter and Alzheimer's disease, Stroke, and White Matter Hyperintensities Volume

| Outcome | Method | Beta | se | P |
| --- | --- | --- | --- | --- |
| Alzheimer's disease | |  |  |  |
|  | MR Egger | -0.001 | 0.002 | 0.792 |
|  | Weighted median | 0.001 | 0.001 | 0.200 |
|  | Inverse variance weighted | 0.001 | 0.001 | 0.170 |
|  | Simple mode | 0.001 | 0.001 | 0.483 |
|  | Weighted mode | 0.001 | 0.001 | 0.356 |
| Stroke |  |  |  |  |
|  | MR Egger | -0.003 | 0.006 | 0.642 |
|  | Weighted median | -0.006 | 0.003 | 0.066 |
|  | Inverse variance weighted | -0.005 | 0.002 | 0.027 |
|  | Simple mode | -0.006 | 0.004 | 0.134 |
|  | Weighted mode | -0.006 | 0.004 | 0.139 |
| White matter hyperintensities volume | | | |  |
|  | MR Egger | -0.37 | 0.172 | 0.050 |
|  | Weighted median | -0.015 | 0.088 | 0.861 |
|  | Inverse variance weighted | -0.069 | 0.062 | 0.268 |
|  | Simple mode | -0.017 | 0.114 | 0.882 |
|  | Weighted mode | -0.012 | 0.103 | 0.910 |

Supplemental table 12 Multi-Trait Analysis of Anterior Brain Arterial Diameter

| SNP | Allele1 | Allele2 | N | P | beta | se | AF |
| --- | --- | --- | --- | --- | --- | --- | --- |
| 12:78464210 | A | G | 3944 | 8.23E-07 | -0.11 | 0.02 | 0.382 |
| 12:78470607 | T | C | 3944 | 1.14E-06 | -0.11 | 0.02 | 0.389 |
| 13:19613694 | T | C | 2630 | 1.29E-06 | 0.14 | 0.03 | 0.7328 |
| 13:19611203 | A | G | 2630 | 1.80E-06 | -0.14 | 0.03 | 0.2651 |
| 13:19608989 | A | G | 2630 | 1.87E-06 | -0.14 | 0.03 | 0.2685 |
| 13:19612262 | A | G | 2630 | 1.87E-06 | -0.14 | 0.03 | 0.265 |
| 13:19610505 | T | G | 2630 | 1.91E-06 | -0.14 | 0.03 | 0.2659 |
| 13:19611302 | T | C | 2630 | 2.28E-06 | -0.14 | 0.03 | 0.2675 |
| 13:19612971 | T | C | 2630 | 2.37E-06 | 0.14 | 0.03 | 0.7346 |
| 12:78472112 | A | G | 3944 | 2.41E-06 | 0.10 | 0.02 | 0.6098 |
| 13:19618596 | A | G | 2630 | 2.45E-06 | 0.14 | 0.03 | 0.7331 |
| 9:93492260 | A | C | 3944 | 2.65E-06 | -0.10 | 0.02 | 0.5671 |
| 13:19610179 | A | C | 2630 | 2.69E-06 | 0.14 | 0.03 | 0.7337 |
| 13:19611764 | A | G | 2630 | 2.89E-06 | 0.17 | 0.04 | 0.8451 |
| 13:19602698 | T | G | 2630 | 3.19E-06 | -0.14 | 0.03 | 0.271 |
| 17:30934596 | T | C | 3473 | 3.22E-06 | 0.12 | 0.03 | 0.2323 |
| 13:19607779 | A | G | 2630 | 3.22E-06 | -0.14 | 0.03 | 0.268 |
| 12:49024269 | A | C | 3508 | 3.35E-06 | 0.12 | 0.03 | 0.2442 |
| 12:49026327 | T | G | 3508 | 3.41E-06 | 0.12 | 0.03 | 0.2442 |
| 12:49021292 | T | C | 3508 | 3.47E-06 | 0.12 | 0.03 | 0.2424 |
| 3:39248474 | A | G | 3028 | 4.06E-06 | 0.18 | 0.04 | 0.1056 |
| 6:19970784 | A | G | 3784 | 4.13E-06 | 0.10 | 0.02 | 0.6054 |
| 15:63127774 | A | G | 3033 | 4.59E-06 | 0.11 | 0.03 | 0.3743 |
| 10:12977850 | A | G | 3739 | 4.73E-06 | -0.13 | 0.03 | 0.8089 |

The statistics are based on Allele1. Allele1 indicates effect allele, allele2 is another allele. AF: allele 1 frequency.

Supplemental table 13 Multi-Trait Analysis of Posterior Brain Arterial Diameter

| SNP | Allele1 | Allele2 | N | P | beta | se | AF |
| --- | --- | --- | --- | --- | --- | --- | --- |
| 16:89948397 | A | G | 3944 | 2.74E-07 | 0.12 | 0.02 | 0.2883 |
| 16:89949033 | T | C | 3944 | 3.60E-07 | -0.12 | 0.02 | 0.7116 |
| 12:61545903 | T | C | 3944 | 4.23E-07 | 0.11 | 0.02 | 0.3465 |
| 12:61544837 | A | C | 3944 | 4.45E-07 | 0.11 | 0.02 | 0.3521 |
| 12:61545307 | T | C | 3944 | 4.45E-07 | 0.11 | 0.02 | 0.3522 |
| 12:61545307 | T | C | 3944 | 4.45E-07 | 0.11 | 0.02 | 0.3522 |
| 12:61545814 | T | C | 3944 | 4.45E-07 | 0.11 | 0.02 | 0.3522 |
| 12:61547083 | A | G | 3944 | 5.26E-07 | -0.11 | 0.02 | 0.6518 |
| 12:61543373 | T | G | 3944 | 5.26E-07 | -0.11 | 0.02 | 0.6465 |
| 16:89939327 | A | G | 3944 | 5.27E-07 | 0.12 | 0.02 | 0.2883 |
| 12:61543784 | A | G | 3944 | 5.37E-07 | -0.11 | 0.02 | 0.6477 |
| 12:61545565 | A | G | 3944 | 5.40E-07 | 0.11 | 0.02 | 0.3503 |
| 12:61542938 | T | G | 3944 | 5.42E-07 | -0.11 | 0.02 | 0.6464 |
| 12:61543904 | A | G | 3944 | 5.54E-07 | 0.11 | 0.02 | 0.3533 |
| 12:61542496 | A | G | 3944 | 5.67E-07 | -0.11 | 0.02 | 0.646 |
| 12:61492205 | A | C | 3784 | 7.08E-07 | 0.11 | 0.02 | 0.3533 |
| 12:61492205 | A | C | 3784 | 7.08E-07 | 0.11 | 0.02 | 0.3533 |
| 12:61494361 | T | G | 3944 | 7.34E-07 | -0.11 | 0.02 | 0.6545 |
| 12:61519686 | T | G | 3944 | 1.05E-06 | -0.11 | 0.02 | 0.6539 |
| 12:61509009 | A | G | 3944 | 1.07E-06 | -0.11 | 0.02 | 0.6538 |
| 12:61514052 | A | G | 3944 | 1.07E-06 | 0.11 | 0.02 | 0.3461 |
| 5:16870820 | A | G | 2692 | 1.07E-06 | 0.14 | 0.03 | 0.3132 |
| 12:61491479 | A | G | 3944 | 1.10E-06 | 0.11 | 0.02 | 0.3449 |
| 12:61536342 | T | C | 3944 | 1.13E-06 | -0.11 | 0.02 | 0.6456 |
| 12:61473705 | A | G | 3654 | 1.15E-06 | 0.11 | 0.02 | 0.3934 |
| 12:61496705 | T | C | 3944 | 1.52E-06 | 0.11 | 0.02 | 0.3464 |
| 2:76491113 | A | C | 2796 | 1.58E-06 | 0.31 | 0.06 | 0.9594 |
| 6:1022733 | A | G | 3784 | 1.62E-06 | 0.10 | 0.02 | 0.446 |
| 7:130917729 | A | G | 3944 | 1.70E-06 | 0.11 | 0.02 | 0.3412 |
| 12:61473716 | T | C | 3654 | 1.78E-06 | -0.11 | 0.02 | 0.6384 |
| 12:61539252 | A | G | 3944 | 1.85E-06 | -0.11 | 0.02 | 0.6373 |
| 16:89965842 | T | C | 3784 | 1.92E-06 | 0.11 | 0.02 | 0.3177 |
| 12:61470275 | A | G | 3654 | 1.96E-06 | 0.11 | 0.02 | 0.3524 |
| 16:89941153 | T | C | 3944 | 2.04E-06 | -0.11 | 0.02 | 0.7028 |
| 7:130916954 | A | G | 3944 | 2.09E-06 | 0.11 | 0.02 | 0.3401 |
| 5:14032887 | A | G | 3784 | 2.13E-06 | 0.11 | 0.02 | 0.3796 |
| 12:61517617 | A | G | 3944 | 2.30E-06 | -0.11 | 0.02 | 0.6514 |
| 12:61542150 | T | G | 3944 | 2.32E-06 | 0.10 | 0.02 | 0.3666 |
| 2:119819724 | T | C | 3870 | 2.51E-06 | -0.10 | 0.02 | 0.4002 |
| 2:119819515 | A | G | 3870 | 2.74E-06 | -0.10 | 0.02 | 0.4 |
| 12:61544358 | A | G | 3944 | 2.87E-06 | -0.10 | 0.02 | 0.6507 |
| 12:61548180 | A | G | 3944 | 2.91E-06 | 0.10 | 0.02 | 0.3501 |
| 12:61539939 | A | C | 3944 | 2.96E-06 | 0.10 | 0.02 | 0.3568 |
| 12:61470406 | A | C | 3654 | 3.21E-06 | 0.11 | 0.02 | 0.3505 |
| 12:61478204 | T | G | 3944 | 3.25E-06 | 0.10 | 0.02 | 0.3403 |
| 12:61478465 | A | G | 3654 | 3.31E-06 | 0.11 | 0.02 | 0.3467 |
| 12:61478428 | T | C | 3654 | 3.44E-06 | -0.11 | 0.02 | 0.6534 |
| 12:61478404 | T | C | 3654 | 3.53E-06 | -0.11 | 0.02 | 0.6528 |
| 14:61644796 | T | G | 3428 | 3.60E-06 | -0.26 | 0.06 | 0.0423 |
| 12:61485147 | A | G | 3944 | 3.62E-06 | 0.10 | 0.02 | 0.339 |
| 12:61486474 | T | C | 3944 | 3.66E-06 | -0.10 | 0.02 | 0.6615 |
| 12:61458805 | A | G | 3944 | 3.67E-06 | 0.10 | 0.02 | 0.3421 |
| 12:61461260 | T | C | 3944 | 3.84E-06 | -0.10 | 0.02 | 0.6624 |
| 12:61461260 | T | C | 3944 | 3.84E-06 | -0.10 | 0.02 | 0.6624 |
| 16:90021685 | A | G | 3733 | 3.85E-06 | -0.11 | 0.02 | 0.6908 |
| 12:61461901 | T | C | 3944 | 3.91E-06 | 0.10 | 0.02 | 0.3382 |
| 12:61470232 | A | G | 3654 | 4.00E-06 | 0.11 | 0.02 | 0.3768 |
| 4:171583722 | A | G | 3784 | 4.06E-06 | 0.10 | 0.02 | 0.3544 |
| 4:171583317 | T | C | 3784 | 4.12E-06 | 0.10 | 0.02 | 0.3544 |
| 12:128860508 | T | C | 3784 | 4.17E-06 | 0.11 | 0.02 | 0.2934 |
| 12:61457051 | A | C | 3944 | 4.50E-06 | -0.10 | 0.02 | 0.6586 |
| 12:61470192 | T | C | 3944 | 4.51E-06 | 0.10 | 0.02 | 0.3393 |
| 12:61459008 | T | C | 3944 | 4.60E-06 | 0.10 | 0.02 | 0.3417 |
| 16:89967397 | A | C | 3784 | 4.71E-06 | -0.11 | 0.02 | 0.6843 |
| 12:61470354 | A | C | 3654 | 4.73E-06 | 0.11 | 0.02 | 0.3587 |
| 12:61474839 | T | C | 3944 | 4.77E-06 | -0.10 | 0.02 | 0.6607 |
| 12:61477095 | T | C | 3944 | 4.77E-06 | -0.10 | 0.02 | 0.6608 |
| 12:61477618 | T | G | 3944 | 4.77E-06 | 0.10 | 0.02 | 0.3391 |
| 12:61477637 | A | G | 3944 | 4.77E-06 | -0.10 | 0.02 | 0.6607 |
| 16:89967217 | T | C | 3784 | 4.81E-06 | -0.11 | 0.02 | 0.6834 |
| 4:113369754 | T | C | 3944 | 4.81E-06 | -0.10 | 0.02 | 0.3298 |
| 12:61475061 | A | C | 3944 | 4.83E-06 | -0.10 | 0.02 | 0.661 |
| 12:61475337 | A | G | 3944 | 4.92E-06 | -0.10 | 0.02 | 0.6609 |
| 12:61476132 | A | G | 3944 | 4.92E-06 | -0.10 | 0.02 | 0.6609 |
| 12:61476675 | A | G | 3944 | 4.92E-06 | -0.10 | 0.02 | 0.6609 |
| 12:61477068 | T | C | 3944 | 4.92E-06 | -0.10 | 0.02 | 0.6609 |
| 4:113354921 | T | C | 3944 | 4.92E-06 | -0.10 | 0.02 | 0.3338 |
| 12:61474412 | A | G | 3944 | 4.99E-06 | -0.10 | 0.02 | 0.6611 |

The statistics are based on Allele1. Allele1 indicates effect allele, allele2 is another allele. AF: allele 1 frequency.

Supplemental Figure 1 Forest plot and PM plot comparing brain arterial diameter estimate for multiple cohorts and meta-analysis estimate for SNP chr10:104840970 (rs7921574) and chr16:89949033 (rs35994878).

| A |  |
| --- | --- |
| 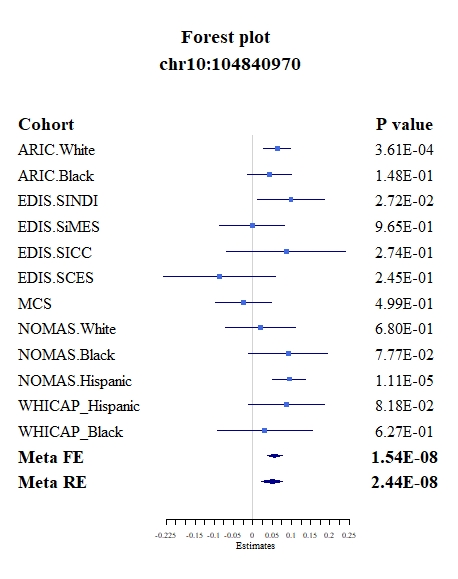 | 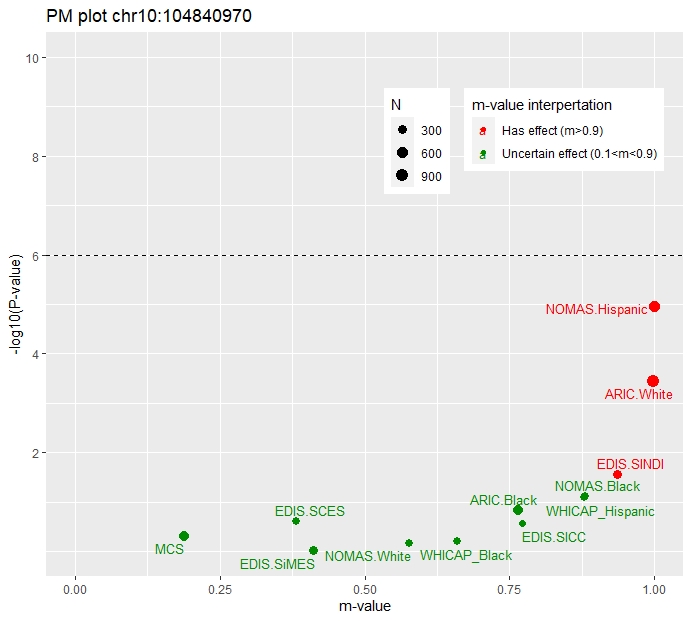 |
| B |  |
| 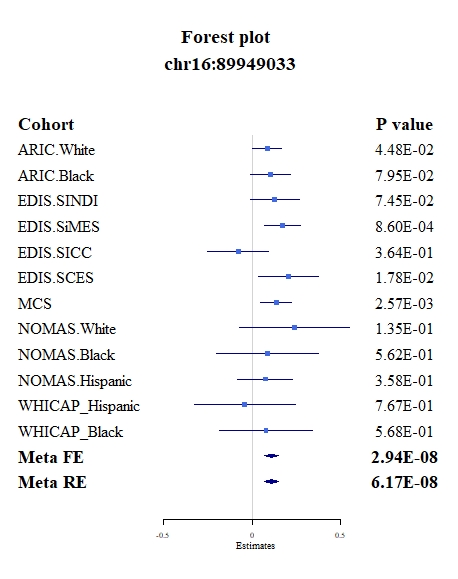 | 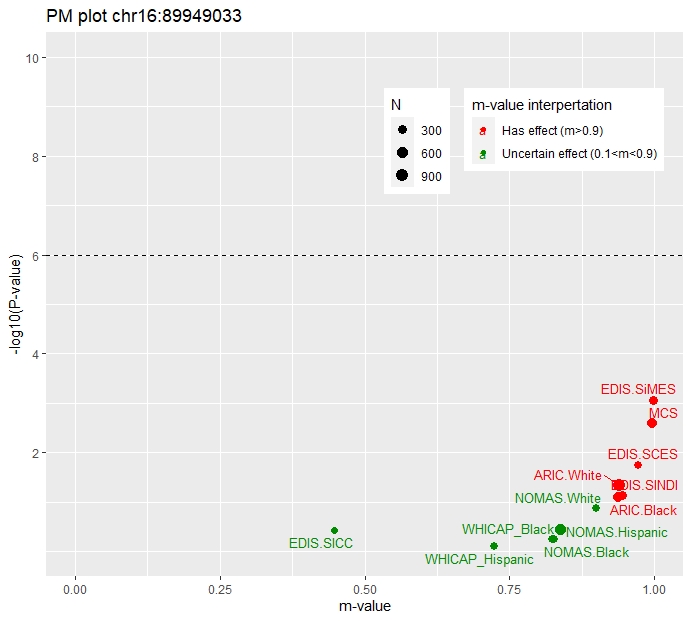 |

Supplemental Figure 2 QQ plot and Manhattan plot of MR-MEGA all study meta-analysis.

| 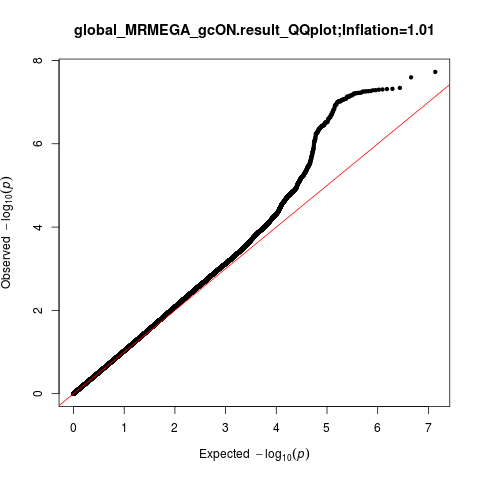 | 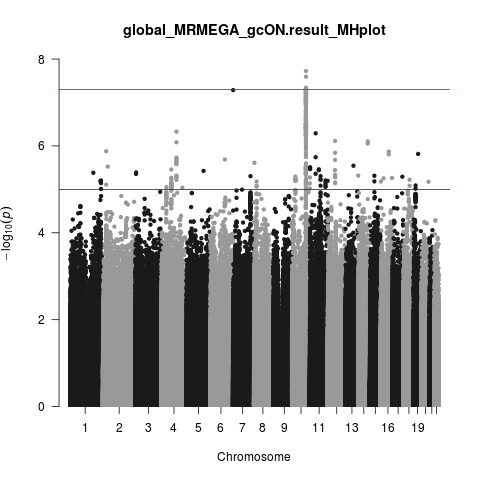 |
| --- | --- |
| 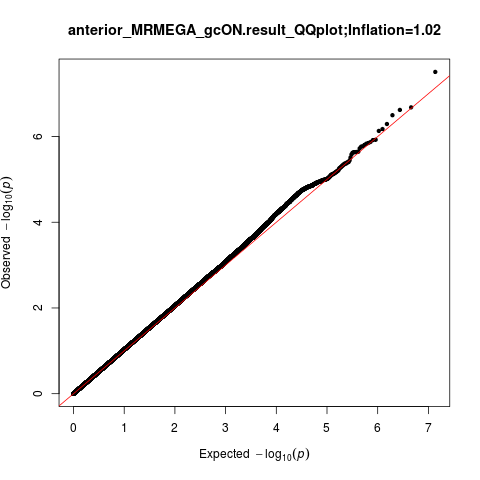 | 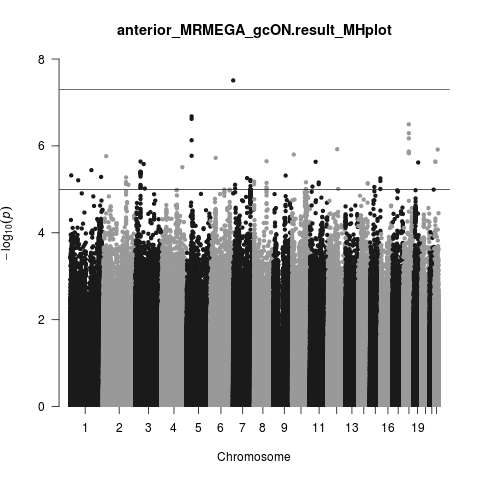 |
| 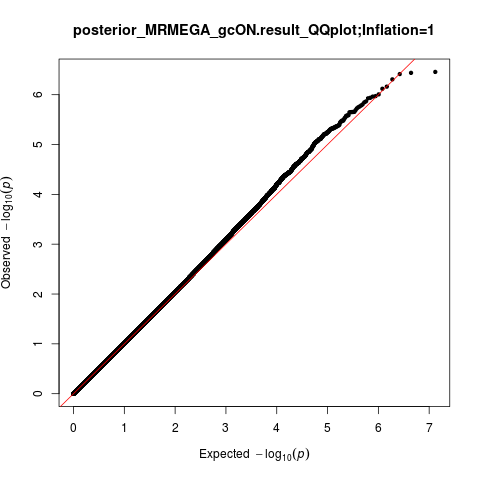 | 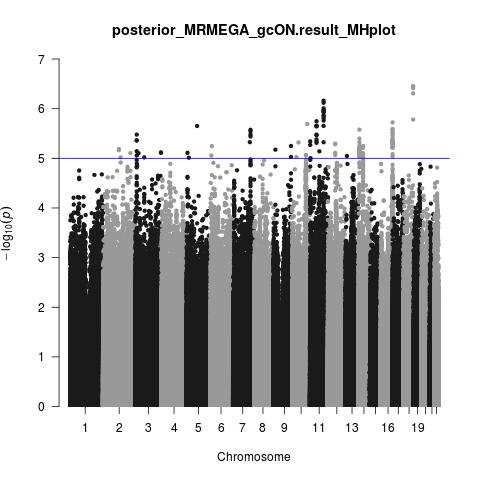 |

Supplemental Figure 3 Locus zoom plot for top SNP rs7921574 in a global brain arterial diameter meta-analysis

| 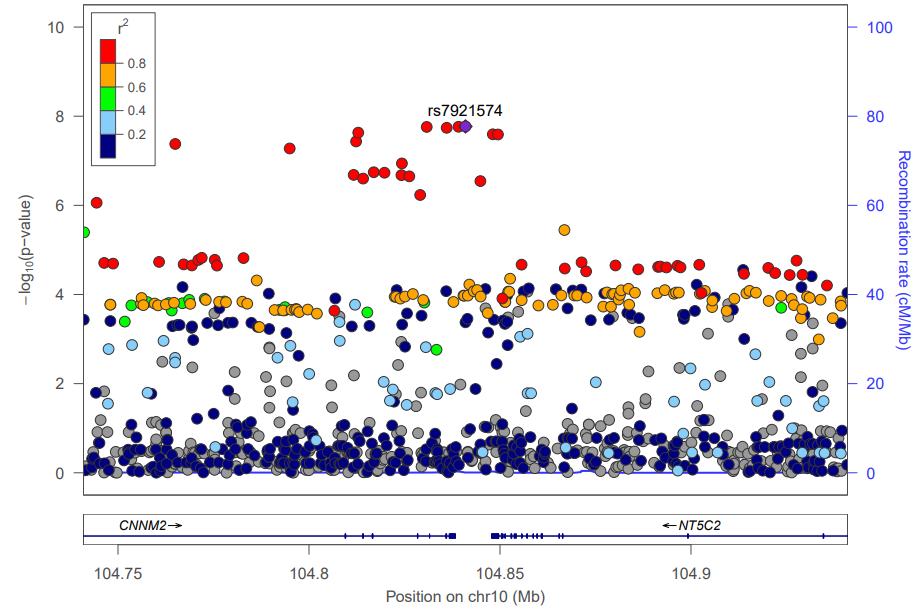  All-study | 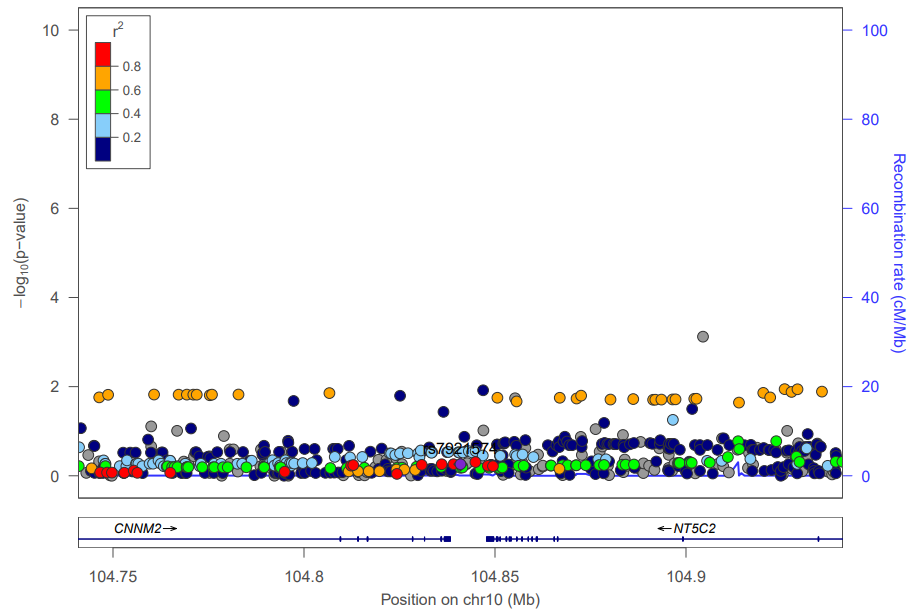  Asian |
| --- | --- |
| 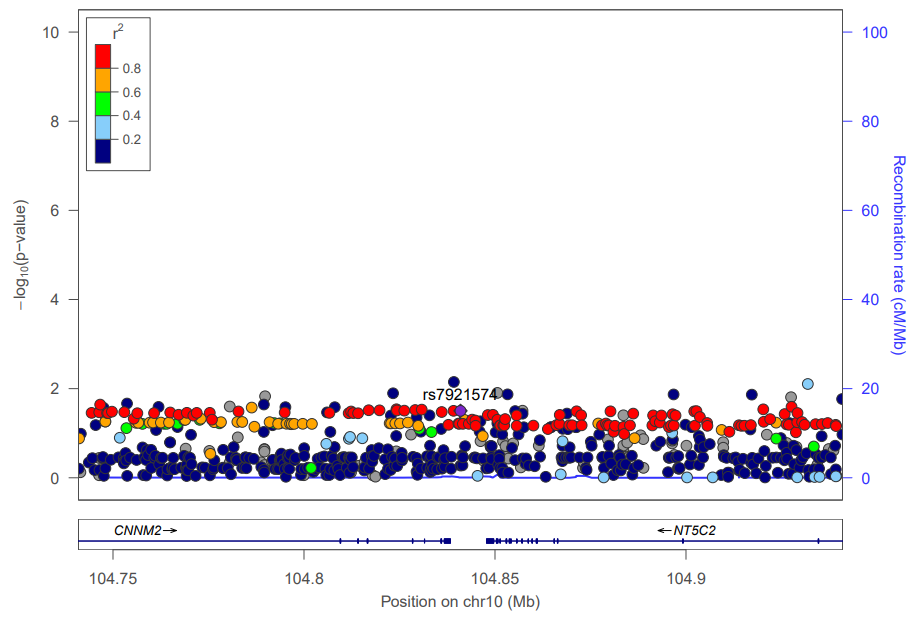  African | 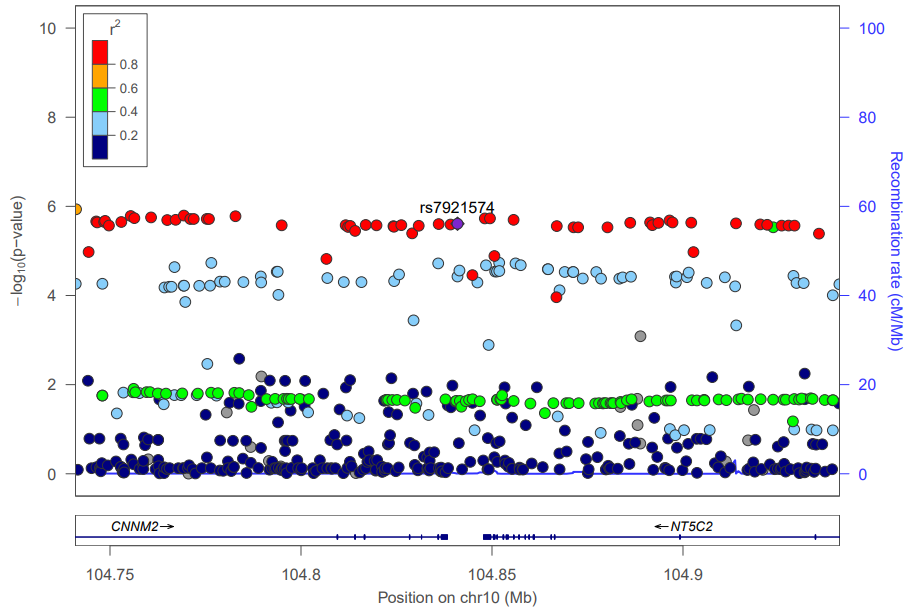  Hispanic |
| 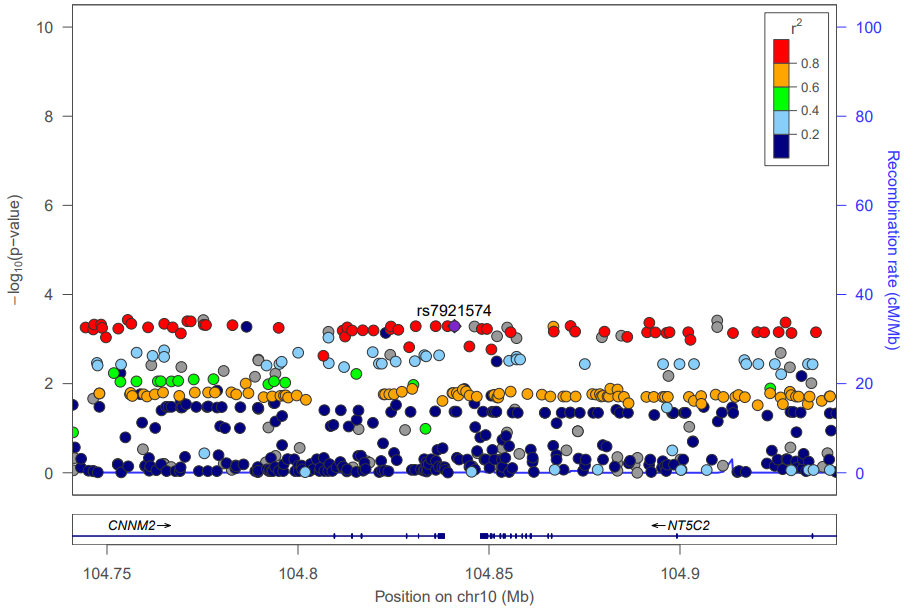  European |  |

Supplemental Figure 4 Locus zoom plot for top SNP rs35994878 in posterior brain arterial diameter meta-analysis

| 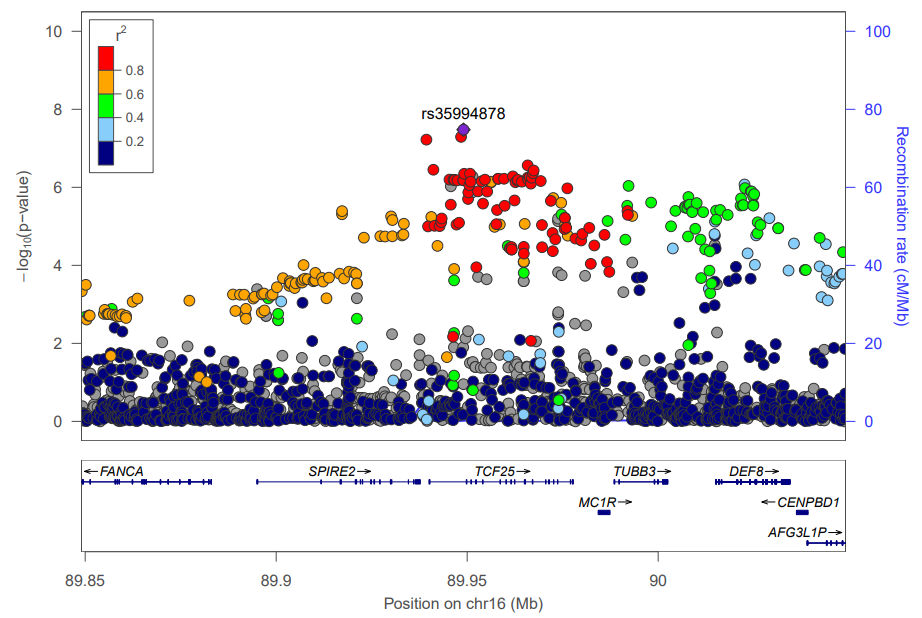  All-study | 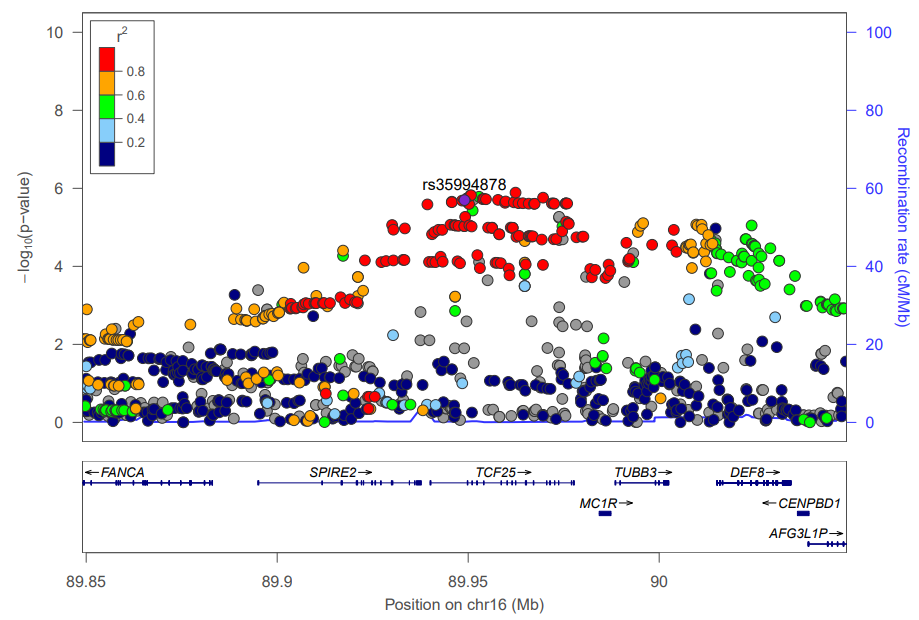  Asian |
| --- | --- |
| 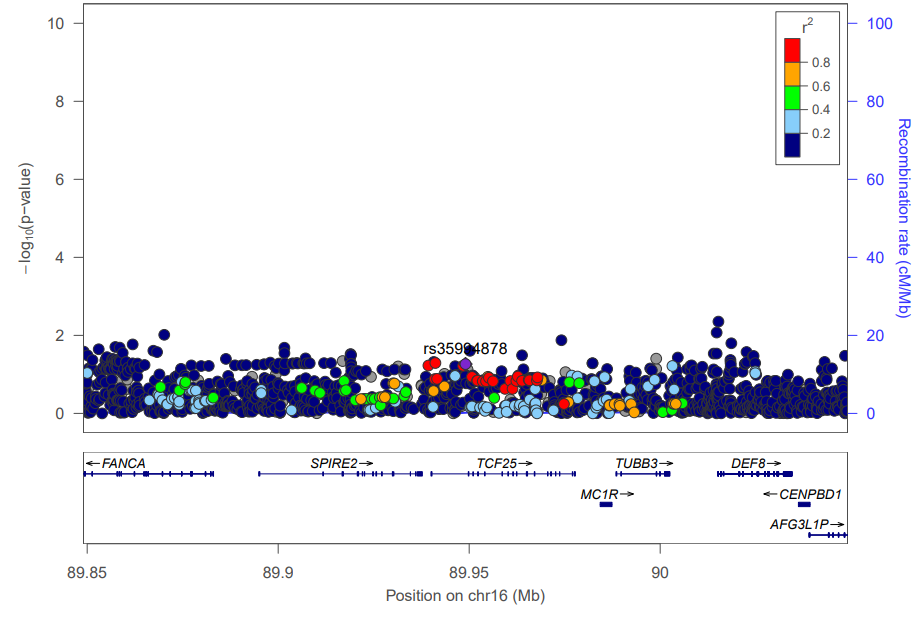  African | 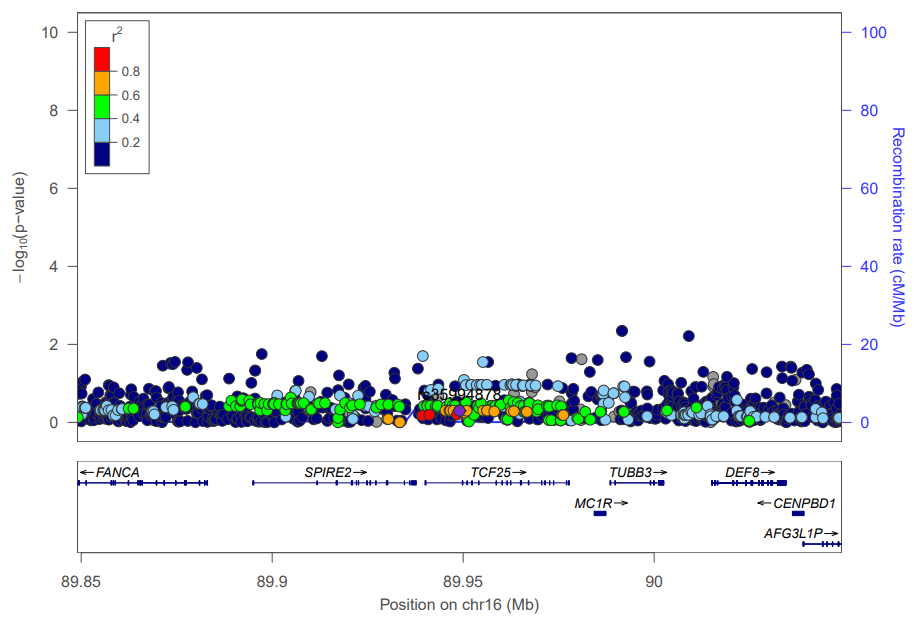  Hispanic |
| 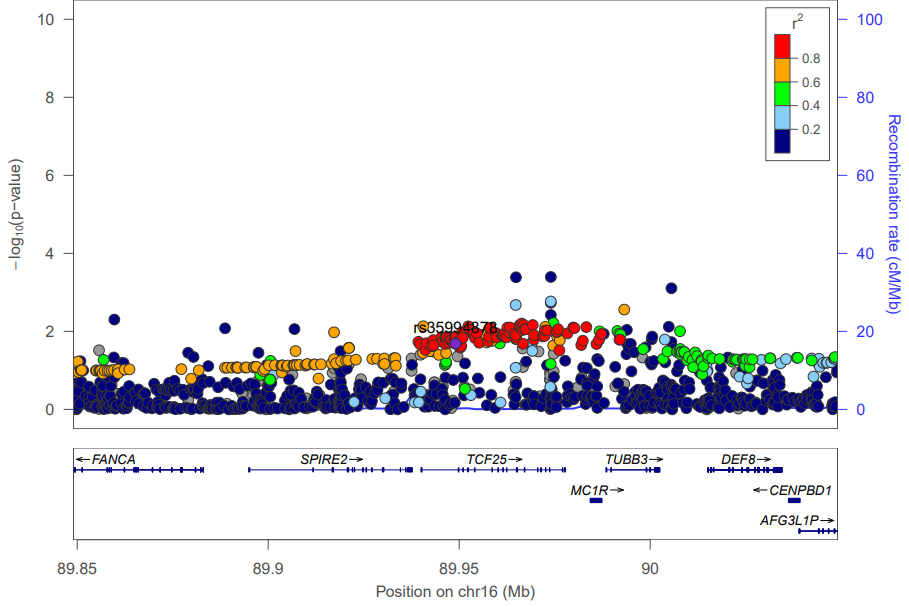European |  |

Supplemental Figure 5 Locus zoom plot for top SNP rs34217429 in anterior brain arterial diameter meta-analysis

| 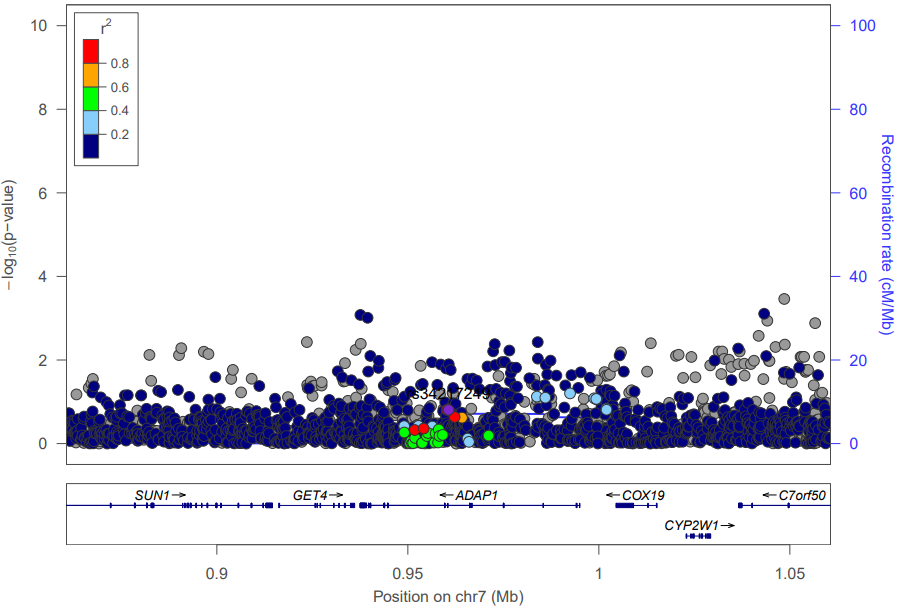  All-study | 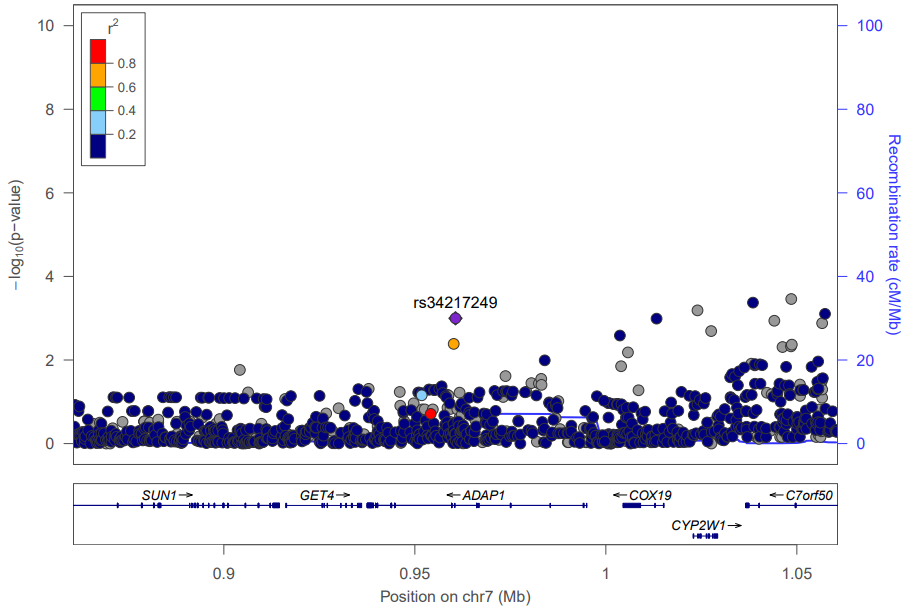  Asian |
| --- | --- |
| 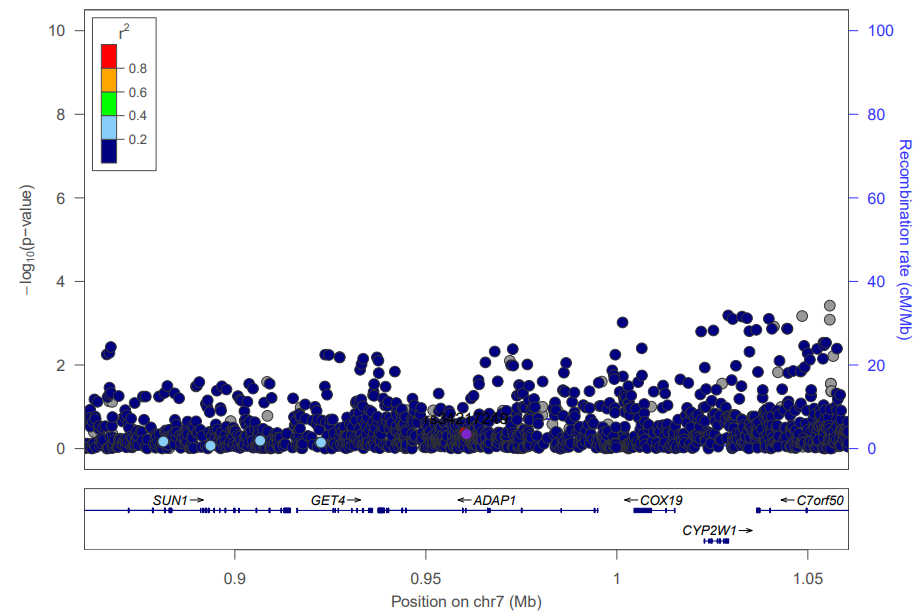  African | 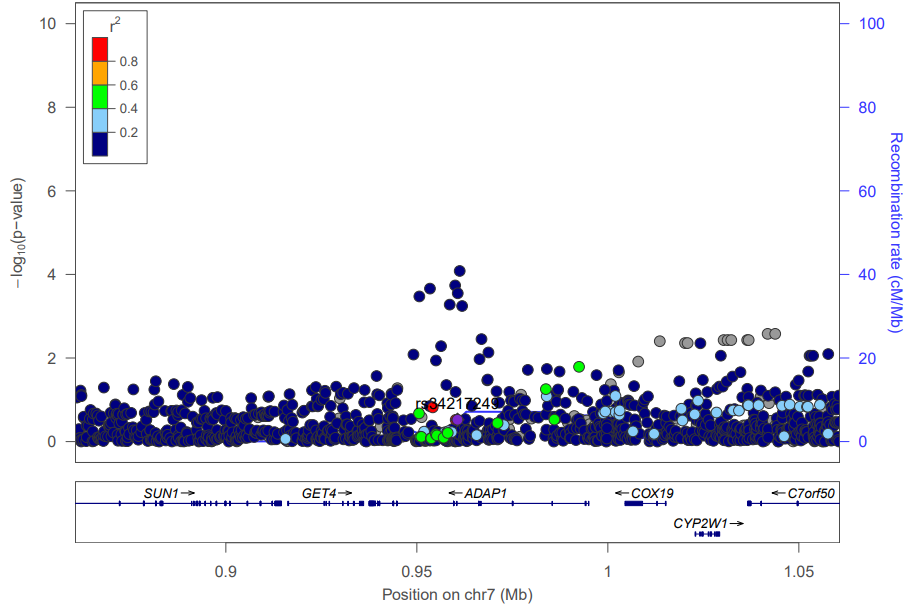  Hispanic |
| 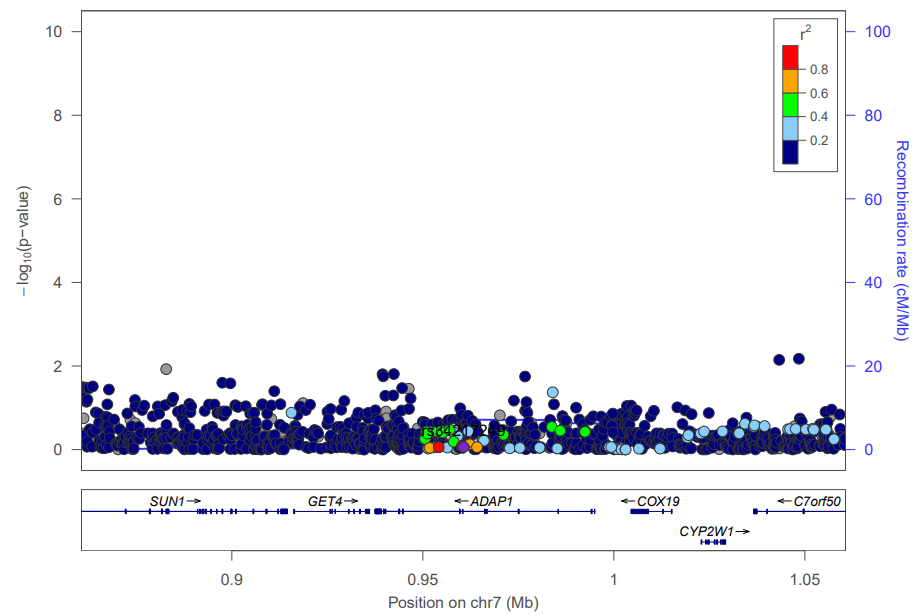  European |  |

Supplemental Figure 6 QQ plot and Manhattan plot of global brain arterial diameter meta-analysis

| A | 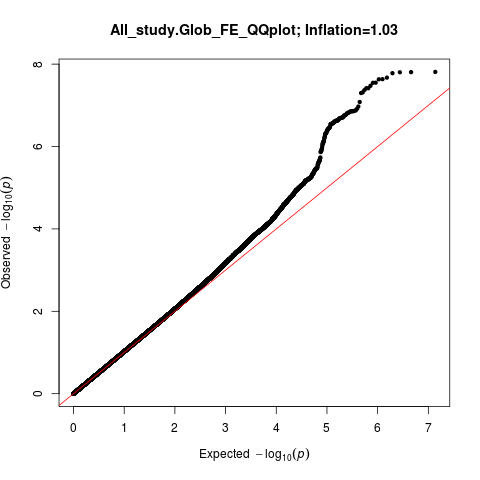 | 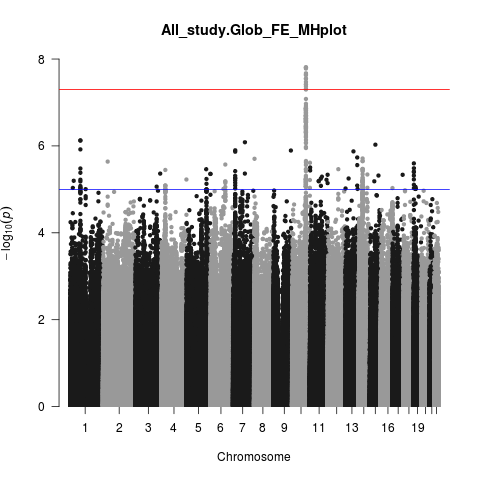 |
| --- | --- | --- |
| B | 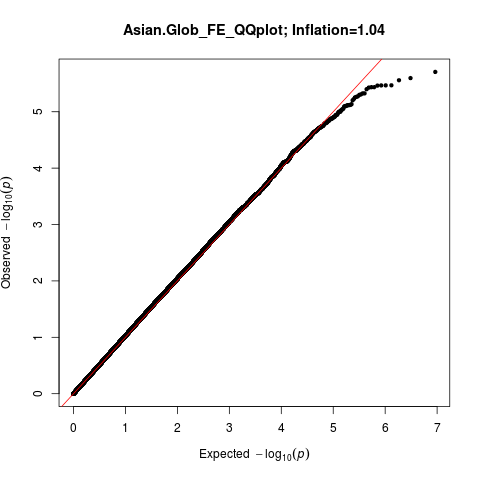 | 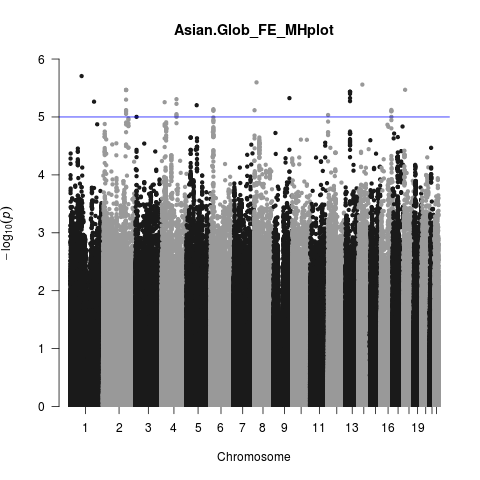 |
| C | 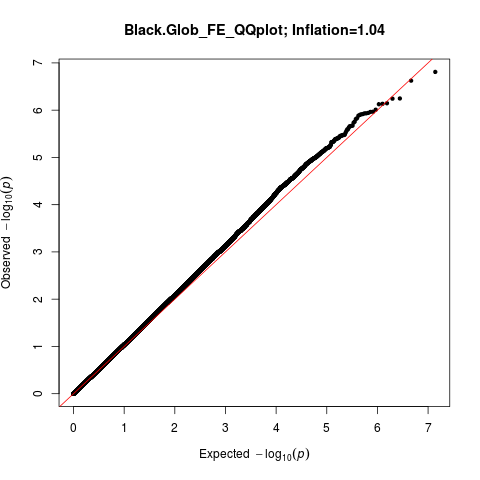 | 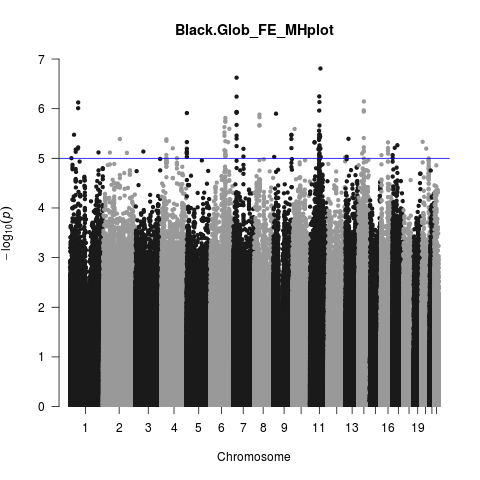 |
| D | 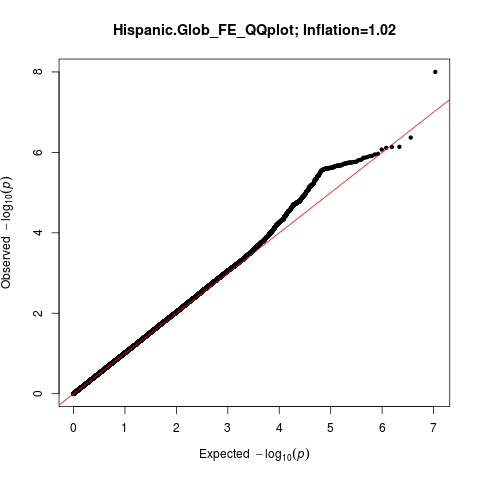 | 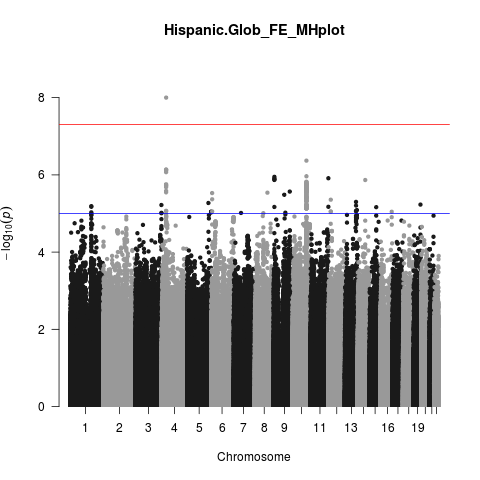 |
| E | 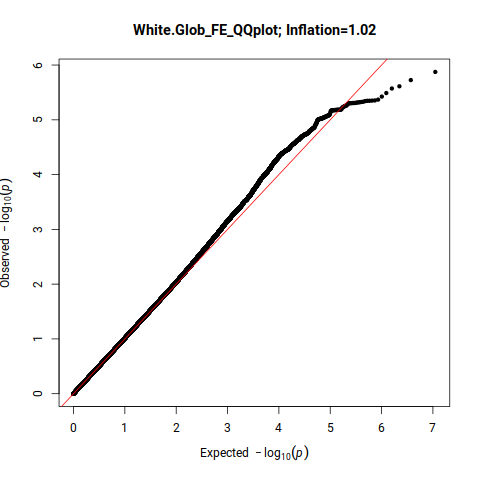 | 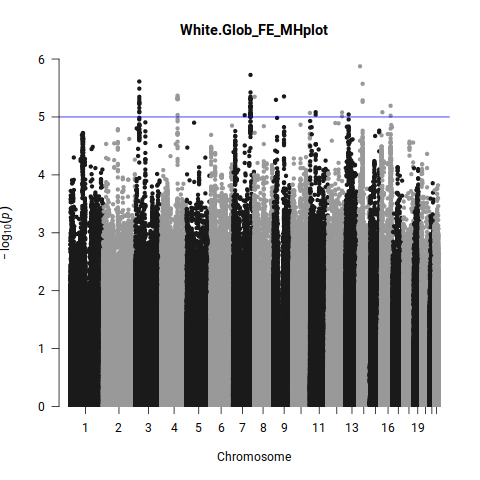 |

Supplemental Figure 7 QQ plot and Manhattan plot of anterior brain arterial diameter meta-analysis

| A | 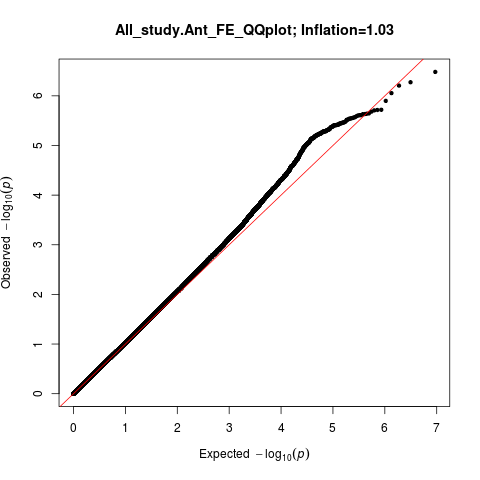 | 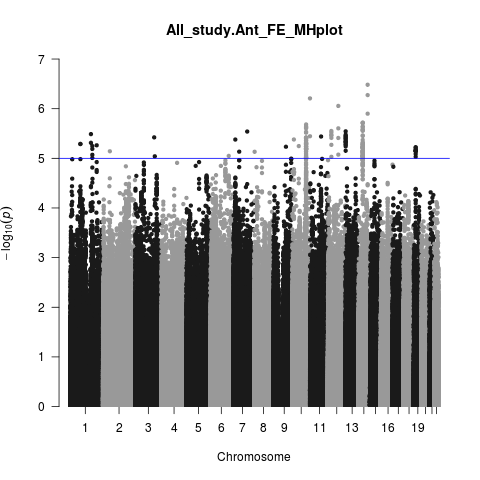 |
| --- | --- | --- |
| B | 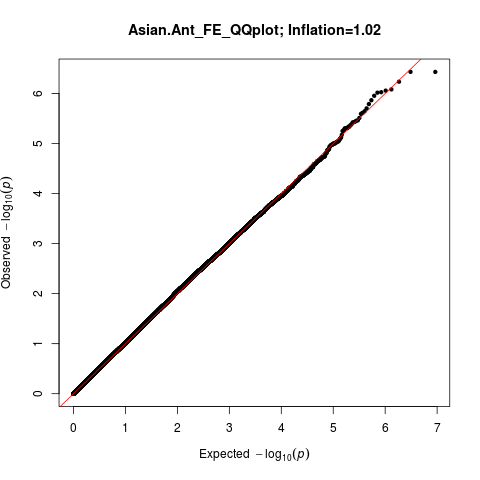 | 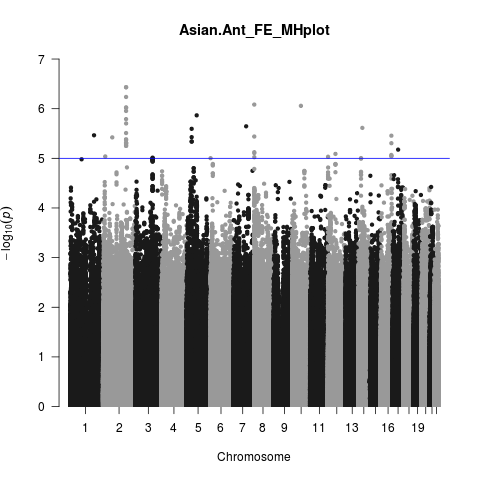 |
| C | 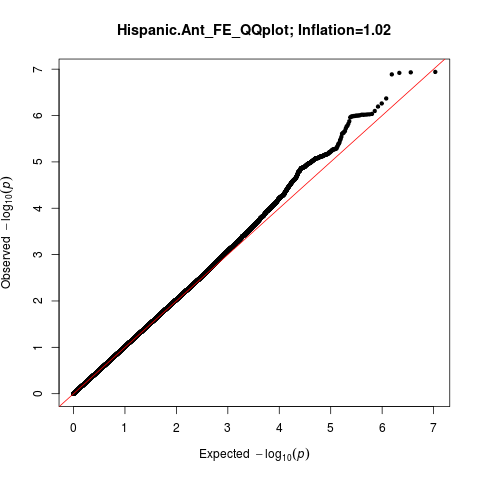 | 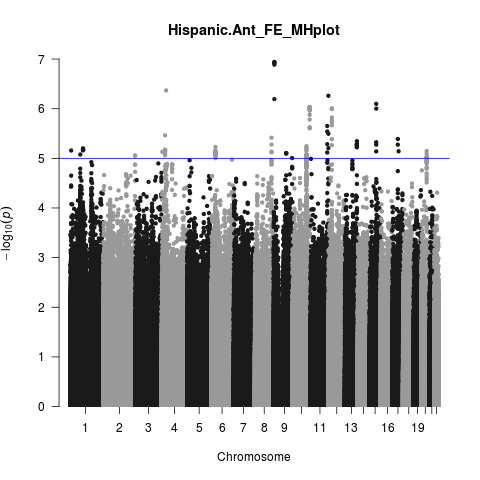 |
| D | 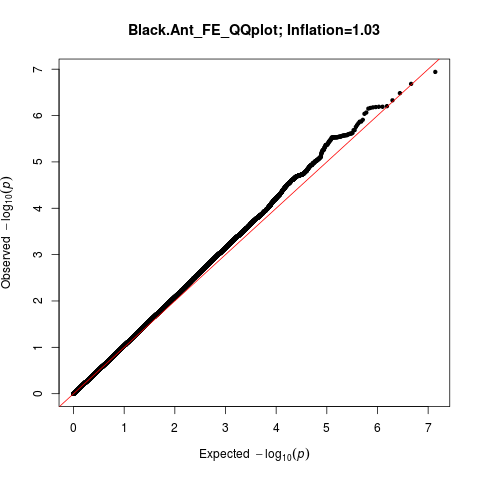 | 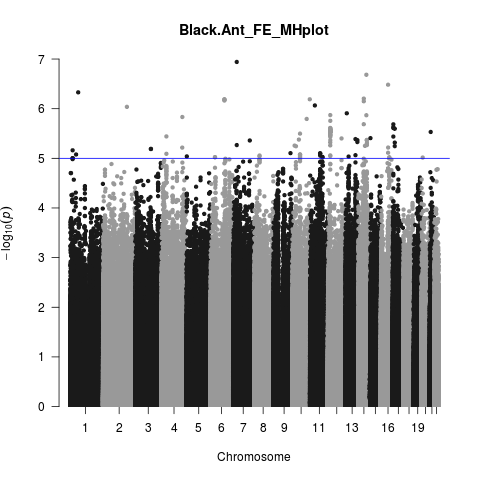 |
| E | 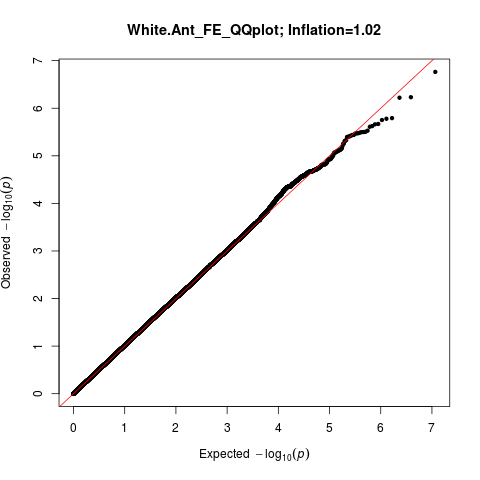 | 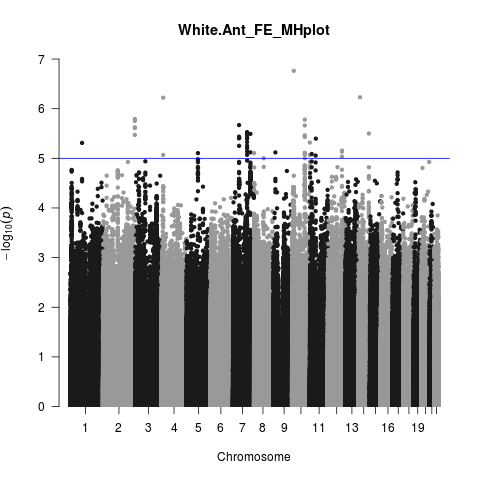 |

Supplemental Figure 8 QQ plot and Manhattan plot of posterior brain arterial diameter meta-analysis

| A | 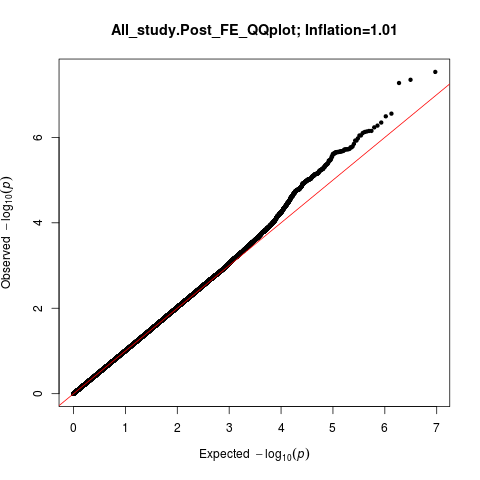 | 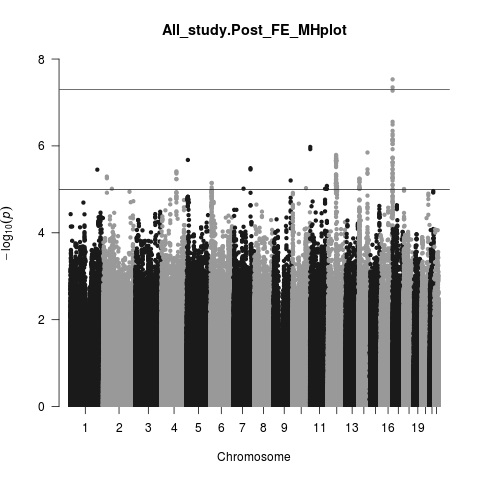 |
| --- | --- | --- |
| B | 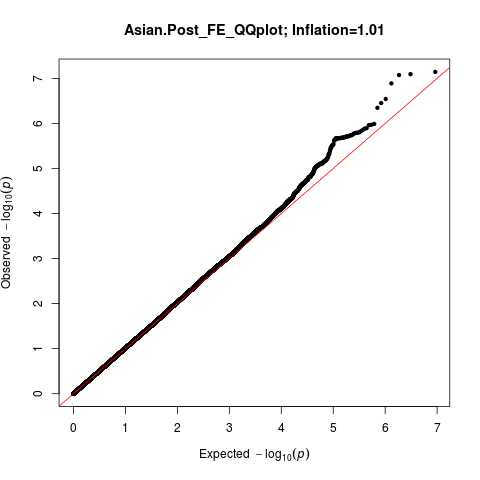 | 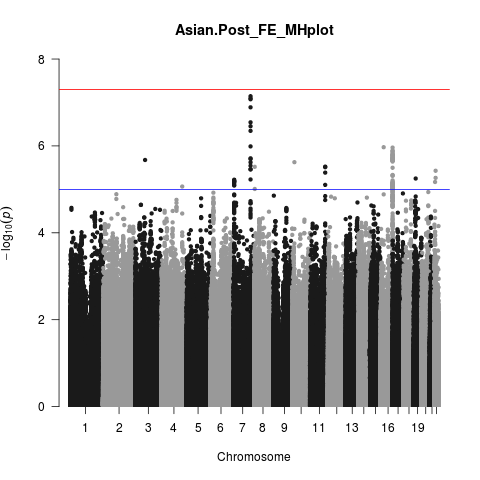 |
| C | 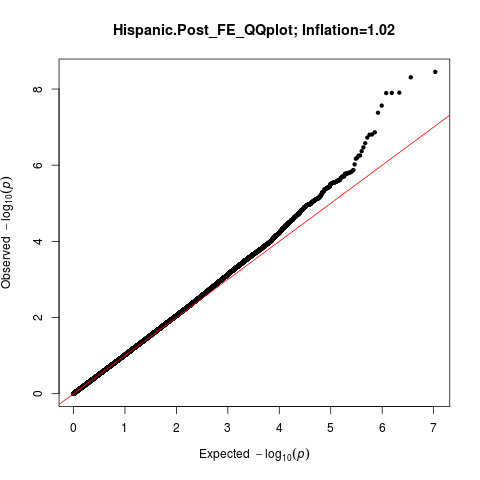 | 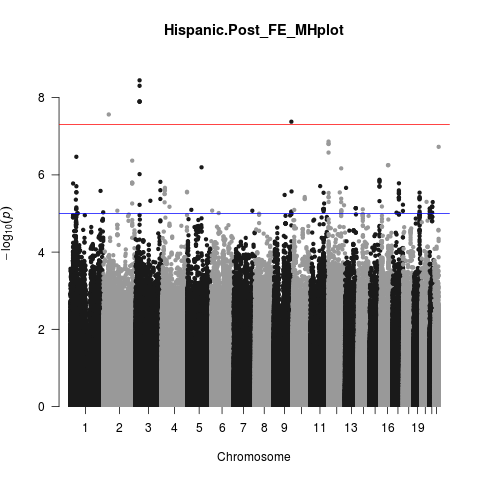 |
| D | 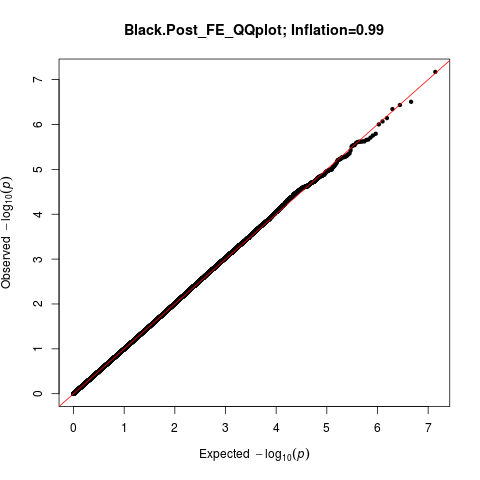 | 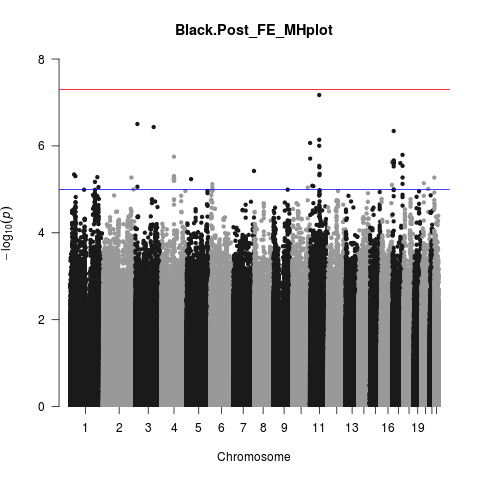 |
| E | 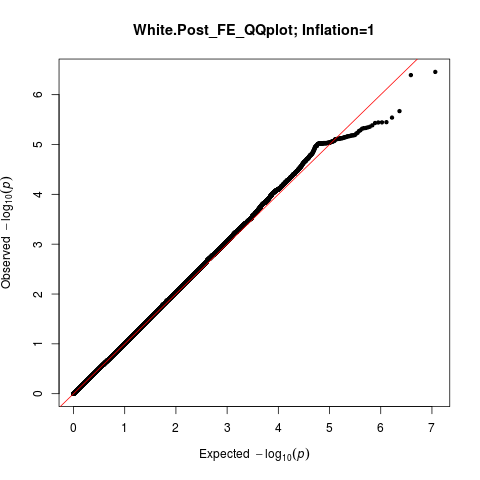 | 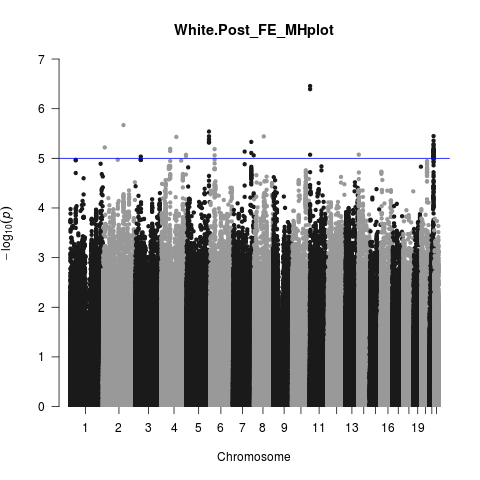 |

Supplemental Figure 9 QQ plot and Manhattan plot of for multivariate analysis of anterior and posterior brain arterial diameter.

| 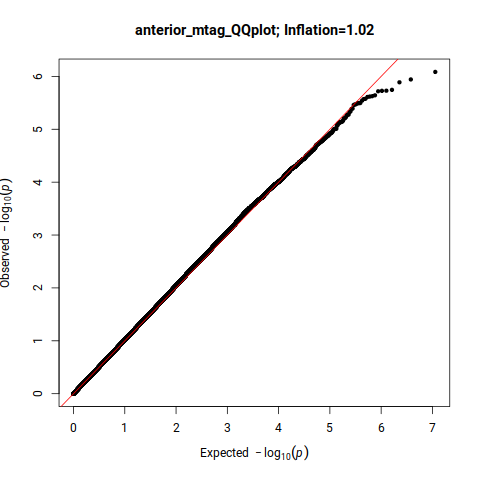 | 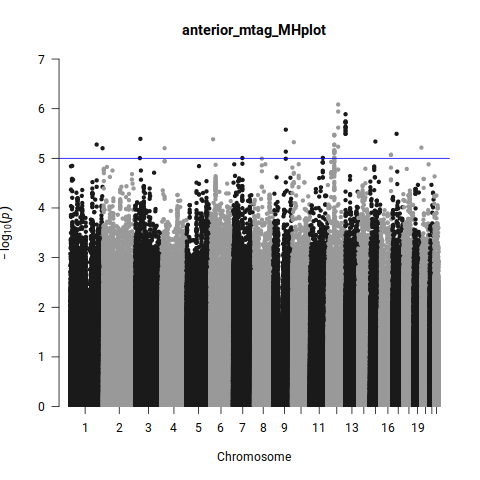 |
| --- | --- |
| 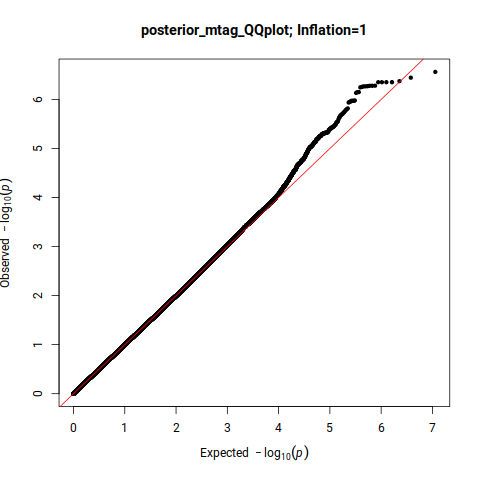 | 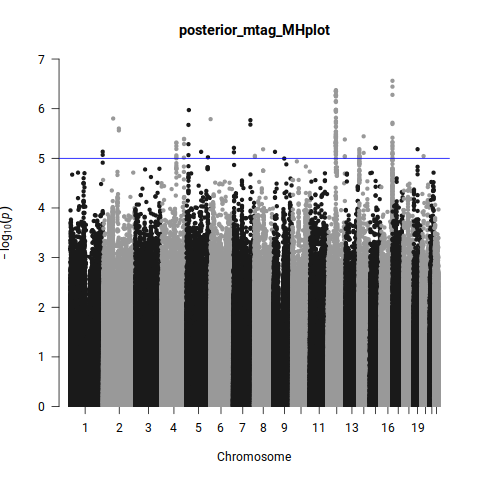 |
